# Weight trends amongst adults with diabetes or hypertension during the COVID-19 pandemic: an observational study using OpenSAFELY

**DOI:** 10.1101/2023.12.17.23300072

**Authors:** Miriam Samuel, Robin Y Park, Sophie V Eastwood, Fabiola Eto, Caroline E Morton, Daniel Stow, Sebastian Bacon, Ben Goldacre, Amir Mehrkar, Jessica Morley, Iain Dillingham, Peter Inglesby, William J Hulme, Kamlesh Khunti, Rohini Mathur, Jonathan Valabhji, Brian MacKenna, Sarah Finer, The OpenSAFELY Collaborative

**Affiliations:** Wolfson Institute of Population Health, Queen Mary University of London, London, United Kingdom; Bennett Institute for Applied Data Science, Nuffield Department of Primary Care Health Sciences, University of Oxford, Oxford, UK; University College London, London, United Kingdom; Leicester Diabetes Centre, Leicester General Hospital, University of Leicester, Leicester, UK; Diabetes Research Centre, College of Medicine, Biological Sciences and Psychology, University of Leicester, Leicester, UK; Division of Metabolism, Digestion and Reproduction, Imperial College London, London, UK; NHS England, Wellington House, London, UK

**Keywords:** Type 2 diabetes, Hypertension, Body Mass Index (BMI), Coronavirus 19 (COVID 19), Health inequalities, Primary Health Care

## Abstract

**Aims:** To describe patterns of weight change amongst adults living in England with Type 2 Diabetes (T2D) and/or hypertension during the COVID-19 pandemic.

**Design and Setting:** With the approval of NHS England, we conducted an observational cohort study using the routinely collected health data of approximately 40% of adults living in England, accessed through the OpenSAFELY service inside TPP.

**Method:** We estimated individual rates of weight gain during the pandemic (δ). We then estimated associations between clinical and sociodemographic characteristics and rapid weight gain (>0.5kg/m^2^/year) using multivariable logistic regression.

**Results:** We extracted data on adults with T2D (n=1,231,455, 44% female, 76% white British) or hypertension (n=3,558,405, 50% female, 84% white British). Adults with T2D lost weight overall (median δ = -0.1kg/m^2^/year [IQR: -0.7, 0.4]), however, rapid weight gain was common (20.7%) and associated with sex (male vs female: aOR 0.78[95%CI 0.77, 0.79]); age, older age reduced odds (e.g. 60-69-year-olds vs 18-29-year-olds: aOR 0.66[0.61, 0.71]); deprivation, (least-deprived-IMD vs most-deprived-IMD: aOR 0.87[0.85, 0.89]); white ethnicity (Black vs White: aOR 0.70[0.69, 0.71]); mental health conditions (e.g. depression: aOR 1.13 [1.12, 1.15]); and diabetes treatment (non-insulin treatment vs no pharmacological treatment: aOR 0.68[0.67, 0.69]). Adults with hypertension maintained stable weight overall (median δ = 0.0kg/m^2^/year [-0.6, 0.5]), however, rapid weight gain was common (24.7%) and associated with similar characteristics as in T2D.

**Conclusion:** Amongst adults living in England with T2D and/or hypertension, rapid pandemic weight gain was more common amongst females, younger adults, those living in more deprived areas, and those with mental health conditions.

**How this fits in:** Previous studies, in the general population, have reported female sex, deprivation and comorbid mental health conditions increased risk of unhealthy weight gain during the pandemic, but it is not clear whether people living with hypertensions and/or type 2 diabetes experienced the same trends.

We found that, during the pandemic, adults with hypertension maintained a stable weight whilst those with type 2 diabetes lost weight overall. However, underlying these overall trends, rapid weight gain was common amongst people with type 2 diabetes (20.7%) or hypertension (24.7%)), with female sex, younger age, deprivation, and comorbid mental health conditions associated with an increased odds of rapid weight gain in both populations.

We have identified clinical and sociodemographic characteristics of individuals with hypertension and/or type 2 diabetes who could benefit from primary care interventions on weight and health behaviours to combat health inequalities in patterns of weight gain that were exacerbated by the pandemic.

## Introduction

Restrictions imposed to reduce COVID-19 transmission resulted in profound societal changes that may have influenced weight-related health behaviours[1–4]. This is of particular relevance amongst adults with type 2 diabetes (T2D) and/or hypertension, conditions which are more common amongst adults with obesity, and interact with obesity to increase the risk of non-communicable disease[5] and serious disease with COVID-19 infection[6].

In addition to absolute weight, rate of weight gain is an independent risk factor for cardiovascular disease[7], and predicts risk of progression to less healthy Body Mass Index (BMI) categories[8]. We previously reported health inequalities in patterns of pandemic weight gain amongst adults living in England, with women, those living in more deprivation and those with mental health conditions at greatest risk of rapid weight gain[9]. Importantly, we found the risk of rapid weight gain was decreased with type 2 diabetes but increased with hypertension[9]. However, whether these trends apply in all socio-demographic groups of adults living with these conditions is unclear. Understanding which subgroups of adults living with type 2 diabetes and/or hypertension were at greatest risk of unhealthy patterns of weight gain during the pandemic will support General Practitioners (GPs) target discussions and interventions on weight and health related behaviours.

We used routinely collected healthcare records to address the following questions amongst adults with T2D and/or hypertension in England during the Covid-19 pandemic: 1) What was the median BMI, and proportion obese, at a population level and within clinical and sociodemographic substrata; 2) What were individual-level patterns of weight change during the pandemic; 3) Which sociodemographic and clinical characteristics were associated with an increased individual-level odds of rapid weight gain (>0.5kg/m^2^/year) during the pandemic.

## Methods

### Data Source

Primary care health records, managed by the GP software provider TPP, were linked, stored and analysed securely within the NHS England COVID-19 OpenSAFELY service inside TPP (OpenSAFELY-TPP), containing pseudonymised data on approximately 40% of the English population, including sociodemographic data, coded diagnoses, medications and physiological parameters. No free text data were included. Detailed pseudonymised patient data is potentially re-identifiable and therefore not shared. An information governance statement is included (Appendix Supplementary 1).

### Data management

Data management was done with Python 3.8 and SQL, and analysis was done using R 4.0. Prevalence counts are rounded to the nearest five to reduce risk of disclosure. All code for data management and analysis, as well as codelists, are shared openly for review and re-use under MIT open license, available at https://github.com/opensafely/BMI-and-Metabolic-Markers.

### Study population

We extracted data on all male and female adults aged 18 to ≤90 years registered with a primary care practice using TPP Electronic Health Record (EHR) software for at least one year prior to 1st March 2022 and identified those who had a coded diagnosis of either T2D and/or hypertension in their healthcare records.

### Study Outcomes: BMI

BMI (weight in kilograms (kg) divided by height in metres squared (m^2^)) is recorded in primary care records during routine health checks, disease monitoring, and when clinically indicated. We extracted BMI data between 1st March 2015 and 1st March 2022. BMI (kg/m^2^) values were obtained either by computing recorded weight and height measurements or, where they were not available, from recorded BMI values present. Weight and BMI measures from before an individual reached 18 years were ignored. Height measurements recorded on individuals aged over 18 years prior to 1st March 2015 were included. We extracted monthly BMI values per individual and where multiple values per calendar month were present, we took the most recent value. Extreme values (BMI<15kg/m^2^ and BMI>65kg/m^2^) were omitted from analyses to censor erroneous results and exclude extremely overweight and underweight individuals whose patterns of weight change may not represent general population trends. We found <0.03% of adults with a BMI value recorded were subsequently excluded from analyses in any given year due to omission of extreme values.

### Cross sectional population level BMI estimates

We used a cross-sectional approach to estimate the population-level median BMI and prevalence of obesity, to give context to the trajectory analysis. BMI values were first categorised by the year in which they were recorded (e.g. March 2021 - March 2022) and individuals were assigned a yearly BMI where data were available. Where multiple values were available in a given year for a single individual, the median was used as the yearly BMI. Due to the reduction in BMI recording during the pandemic, population level descriptive BMI statistics are estimated using the most recent yearly BMI recorded in the 5 years preceding March 2022. Individuals were classified as being obese if they had a BMI >= 30kg/m^2^ (>= 27.5 in Black, Asian or ethnic minority).[10]

### Individual level BMI trajectories during the pandemic

We estimated individual level BMI trajectories as rates of BMI change per year (δ) in kg/m^2^/year. To calculate δ we first categorised recorded BMI values into the following time windows: Period 1 (March 2018 – February 2020) and Period 2 (March 2020 – February 2022) and then, for individuals with BMI data available, randomly selected one BMI value recorded in each of these periods. Amongst adults with a BMI value recorded in each of these periods, we then calculated individual level rates of weight change (δ) between Period 1 and Period 2 assuming a linear trend[8, 9]. (Figure 1, Appendix Supplementary 2). Individuals who were underweight (BMI <18.5kg/m^2^) in Period-1 and those with known cancer were excluded from these analyses as their patterns of weight change were likely to differ from the general population. The most extreme 0.05% of values (a positive or negative change of >6kg/m^2^/year) were censored to reduce the impact of erroneous results. To further contextualise these results, we calculated the rate of weight gain in the prepandemic period using similar methods. (Figure 1, Appendix Supplementary 2).

**Figure 1:**
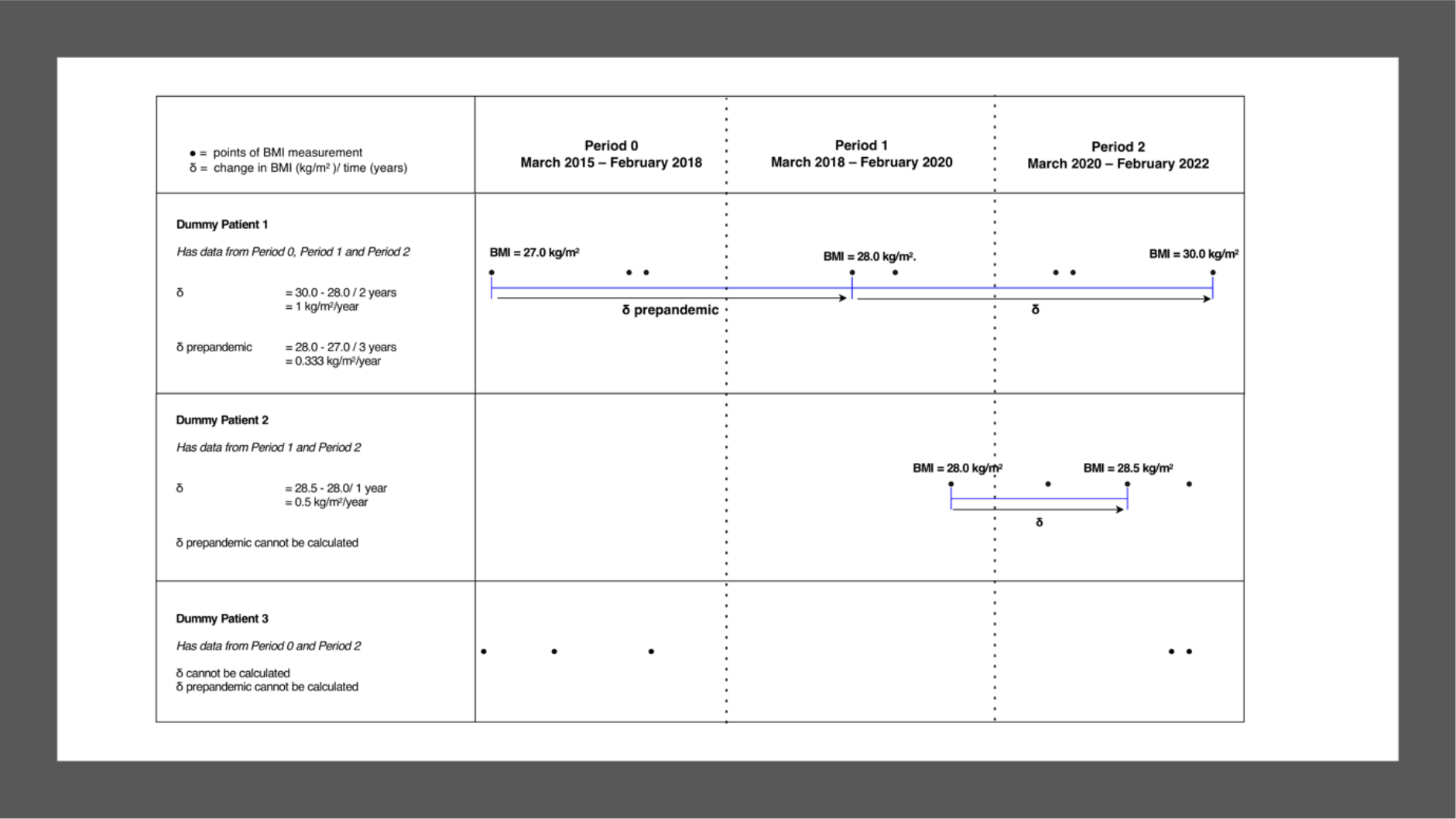
Calculation of rate of Body Mass Index (BMI) change per year (δ) in kg/m^2^/year

Using individual level BMI trajectories, we subsequently categorised adults who gained >0.5kg/m^2^/year as experiencing ‘rapid weight gain’. This rate is independently associated with cardiovascular morbidity [7].

### Covariates

Covariates included age, sex, ethnicity (based on the 2001 UK Census definitions[11]), deprivation (using patient postcode derived Index of Multiple Deprivation(IMD)[12]) and the presence of 14 long-term conditions ever recorded. (Appendix Supplementary 3) Adults with T2D were further classified based on their prescribed diabetes medication regimen into the following three groups: insulin-based regimens (insulin), regimens that did not include insulin (non-insulin), and those who were not prescribed pharmacological treatment (Appendix Supplementary 3).

### Statistical Models

We used complete case analysis, i.e., only included individuals with all baseline covariate data. Complete data were available for age and sex, as these were part of the study inclusion criteria, and long-term conditions (such as depression), which were identified based on the presence/absence of specified clinical codes. Therefore, ethnicity and IMD were the only variables with missing data. The missing at random assumption (required for imputation) was unlikely to hold, e.g., presence of recorded ethnicity is likely to be related to the ethnicity itself. However, the complete case assumption of independence between missingness and outcomes (conditional on the covariates) was more plausible.[13]

We compared the baseline characteristics of the populations with T2D and/or hypertension with and without the outcome (rapid pandemic weight gain). Further comparison of baseline characteristics and outcomes was made between the complete case sample and entire population, including those with missing ethnicity and deprivation data.

Logistic regression was used to explore associations between the sociodemographic and clinical covariates and the odds of rapid weight gain amongst adults with hypertension or T2D. Models were adjusted separately for age, sex, IMD and ethnicity and then in multivariable models adjusted for age and sex; age, sex and IMD; age, sex and ethnicity; and age, sex, IMD and ethnicity.

### Subgroup Analyses

We investigated whether estimated associations between the covariates and extreme acceleration in rate of weight gain persisted in populations stratified by age group (18-39 years, 40-59 years, and 60-79 years), sex, and ethnicity (Black and South Asian).

### Patient and public involvement

OpenSAFELY has a publicly available website through which we invite patients or members of the public to contact us about this study or the broader OpenSAFELY project.

## Results

Data were extracted on 1,231,455 adults with T2D (44% female, 76% white British) and 3,558,405 adults with hypertension (50% female, 84% white British), of which 792,360 adults had both hypertension and T2D and were included in both arms of the analysis (Figure 2 and Table Supplementary 1). Amongst adults with T2D, 64.4% had comorbid hypertension, while 25.7% had comorbid depression. Amongst adults with hypertension, 22.3% had comorbid T2D, while 23.2% had comorbid depression. Few adults with T2D had missing data for ethnicity (3.6%) or IMD (2.0%). Missing data for ethnicity (4.7%) and IMD (1.9%) was also uncommon amongst adults with hypertension (Table Supplementary 2).

**Figure 2:**
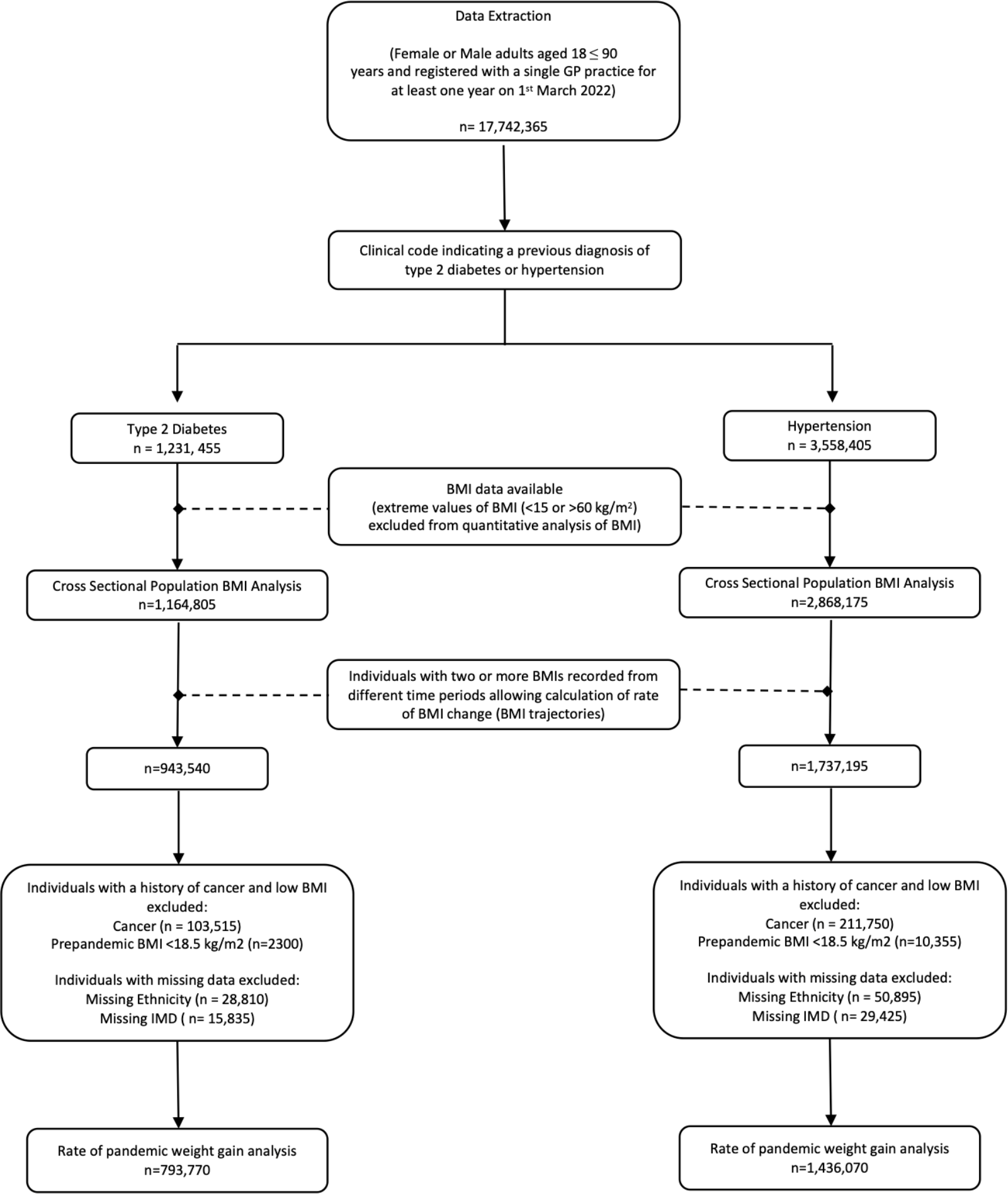
Selection of the study population

### Cross Sectional Population BMI Estimates

Amongst adults with T2D, over half were obese (55.5% obese, median BMI:30.3kg/m^2^ [IQR:26.6, 34.8]) (Table Supplementary 3). BMI was higher amongst: younger adults (18 to 29 year olds: 35.2kg/m^2^ [29.7, 41.1]); women (31.1kg/m^2^ [26.9, 36.2]); those in the most deprived IMD quintiles (31.0kg/m^2^ [27.1, 35.8]); those with mental health conditions including depression (31.6kg/m^2^ [27.6 to 36.5]); and those taking insulin (31.3kg/m^2^ [27.4, 36.0]). White British adults living with T2D had the highest BMI (30.8kg/m^2^ [27.1, 35.3]), followed by those with a mixed White/Black Caribbean ethnicity (30.7kg/m^2^ [26.8, 35.5]) (Figure 3).

**Figure 3:**
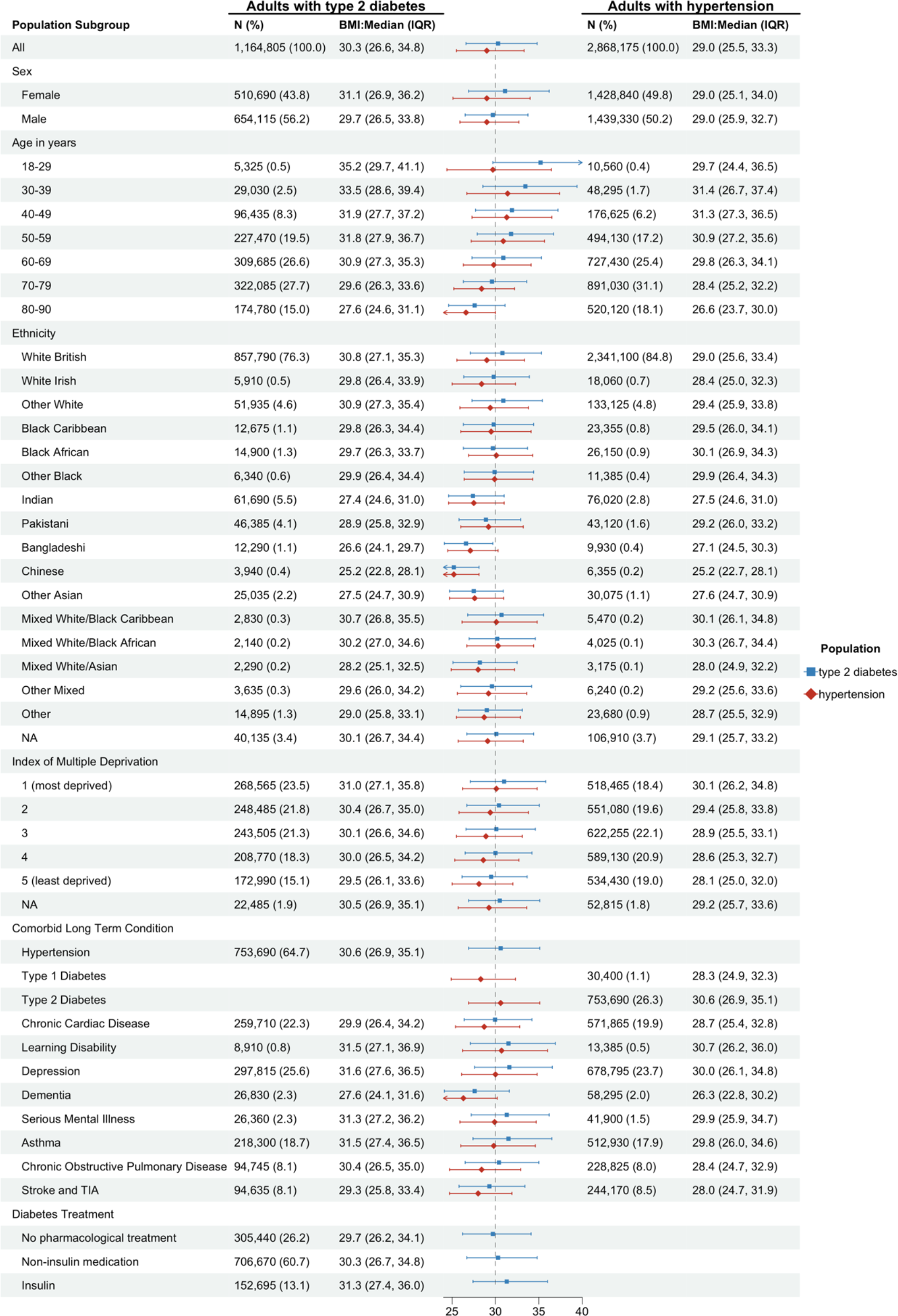
Population level Body Mass Index (BMI) estimates amongst people with type 2 diabetes and/or hypertension. BMI (in kilograms/metre^2^) were calculated from recorded weight (kg) and height (m) measures, or directly from recorded BMI values. N (%): Total number (N) and percentage (%) from each population strata contributing to the analysis. IQR: Interquartile Range. TIA: Transient Ischaemic Attack.

Amongst adults with hypertension, just under half were obese (45.2%, Table Supplementary 3), BMI was higher amongst those living in the most deprived IMD quintile (30.1kg/m^2^ [26.2, 34.8]), and those living with comorbidities including T2D (30.6kg/m^2^ [26.9, 35.1]) and mental health conditions (e.g. depression: 30.0kg/m^2^ [26.1,34.8]). There was no difference in BMI between males and females with hypertension (Figure 3).

### Individuals level BMI trajectories (δ) during the pandemic

During the pandemic, adults with T2D lost weight overall (median δ = -0.1kg/m^2^/year [IQR: -0.7, 0.4]). Only adults taking insulin and those receiving no pharmacological treatment for diabetes maintained a stable weight on average (Figure 4). There was a wide distribution in individual level BMI trajectories, with 25% of adults with T2D gaining at least 0.4kg/m^2^/year (Figure 4). In comparison, the distribution of individual level BMI trajectories was narrower before the onset of the pandemic, although the median rate of weight change was the same (median δ-prepandemic = -0.1kg/m^2^/year [IQR: -0.6, 0.3]) (Figure Supplementary 1).

**Figure 4.**
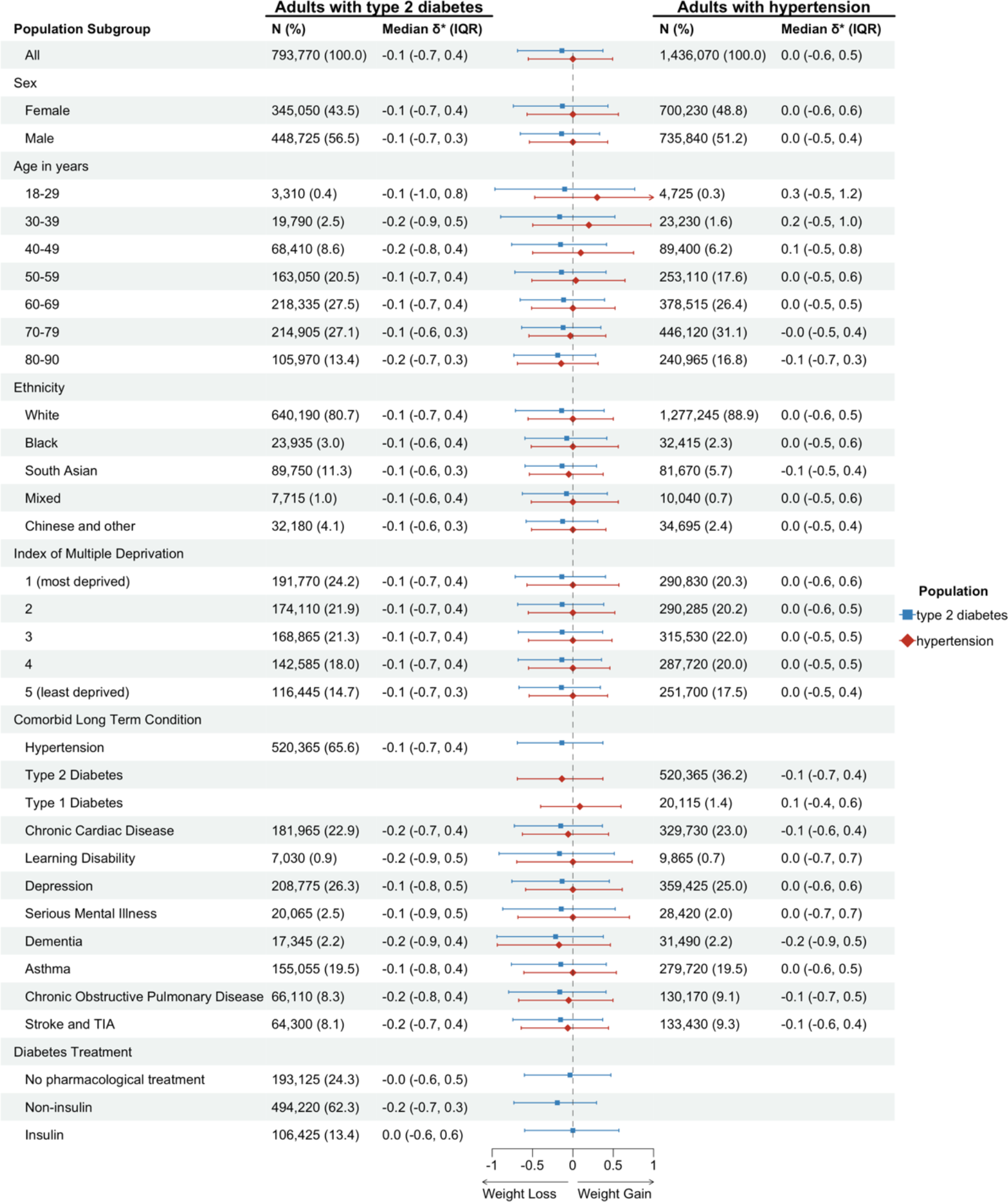
Average rate of weight gain (δ) during the COVID-19 pandemic amongst adults with diabetes and/or hypertension. δ: individual level rate of weight change (in kilograms/metre^2^/year) calculated from data recorded in routine health care records. N (%): Total number (N) and percentage (%) from each population strata contributing to the analysis. IQR: Interquartile Range. TIA: Transient Ischaemic Attack.

Adults living with hypertension maintained a stable weight overall, but there was a wide distribution in individual level BMI changes, with 25% of individuals gaining at least 0.5kg/m^2^/year (median δ = 0.0kg/m^2^/year [-0.6, 0.5]) (Figure 4). Amongst adults with hypertension those aged under 50 years old and those with comorbid type 1 diabetes gained weight, whilst those with comorbid T2D lost weight overall (Figure 4). As seen amongst adults with T2D, there was a narrower distribution of BMI trajectories before the onset of the pandemic, but overall adults with hypertension maintained a stable weight during this time (median δ-prepandemic = 0.0kg/m^2^/year [-0.5, 0.4]) (Figure Supplementary 1).

### Rapid weight gain (δ >0.5kg/m^2^/year) during the pandemic

During the pandemic, 20.7% of adults living with T2D and 24.7% of adults living with hypertension gained weight rapidly (Figure 5).

**Figure 5.**
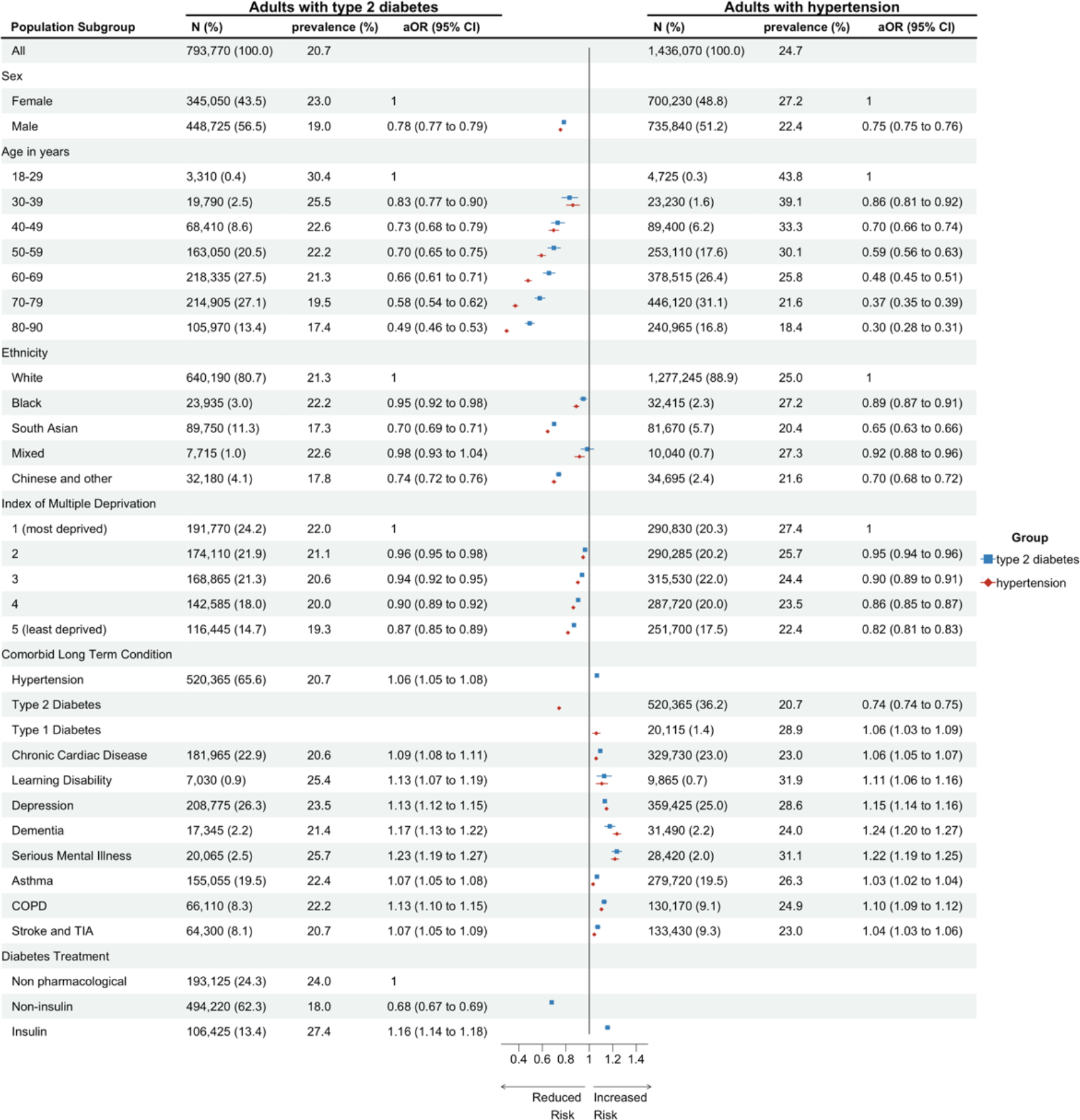
Prevalence and adjusted odds of rapid weight gain (>0.5kg/m^2^/year) amongst adults living in England during the COVID-19 pandemic. N (%): Total number (N) and percentage (%) from each population strata contributing to the analysis. Prevalence (%): percentage who gained weight rapidly during the COVID-19 pandemic. COPD: Chronic Obstructive Pulmonary Disease. TIA: Transient Ischaemic Attack.

Amongst adults with T2D, sociodemographic factors associated with rapid weight gain included: sex, the prevalence was highest amongst women (23%), while male sex reduced the likelihood (male vs female: aOR 0.78[95%CI 0.77 to 0.79]); age, the prevalence was highest amongst 18-29 year olds (30.4%), while older age reduced the likelihood (60-69-year-olds vs 18-29-year-olds: aOR 0.66[0.61, 0.71]); deprivation, prevalence was highest amongst adults living in the most deprived IMD quintile (IMD-1:22.0%), while living in less deprived areas reduced the likelihood (IMD-5 vs IMD-1: aOR 0.87[0.85, 0.89]); and ethnicity, a slightly higher proportion of Black adults than White adults gained weight rapidly (22.2% vs 21.3%), but Black adults had a lower likelihood of rapid weight gain in analyses adjusted for age, gender and deprivation (Black vs White: aOR 0.95[0.92, 0.98])(Figure 5, Table Supplementary 4) Individuals with comorbid conditions had an increased likelihood of rapid weight gain, with mental health conditions associated with the greatest estimated effect (e.g., depression: aOR 1.13 [1.12, 1.15]). The prescribed diabetes treatment was also associated with likelihood of rapid weight gain: adults prescribed non-insulin treatment had a reduced likelihood compared to those who had no pharmacological treatment (aOR 0.68[0.67, 0.69]), whilst those prescribed insulin had an increased likelihood (aOR 1.16[1.14, 1.18]). These sociodemographic and clinical associations were maintained in a multivariate model mutually adjusted for age, sex, IMD, ethnicity and diabetes medication (Figure Supplementary 2). Similar factors increased the adjusted odds of rapid weight gain amongst adults living with hypertension (Figure 5). But, consistent with findings in T2D, amongst adults with hypertension, those with comorbid T2D had lower likelihood of rapid weight gain compared to those without (aOR 0.74[0.74, 0.75]).

To contextualise these findings, we report that before the onset of the pandemic, the prevalence of rapid weight gain was lower amongst adults with T2D (19.2%) and hypertension (21.8%), however, patterns of associations with the clinical and sociodemographic characteristics were similar. (Figure Supplementary 3)

### Subgroup Analyses

Trends seen in the whole population of adults living with T2D and/or hypertension were maintained in the analyses stratified by gender, ethnicity and age. (Figure Supplementary 4-9).

## Discussion

### Summary

We present the first large-scale analysis of weight trends amongst adults living with T2D and/or hypertension during the pandemic. Obesity was common amongst adults living with these conditions. However, people living with T2D tended to lose weight during the pandemic, whilst those living with hypertension maintained a stable weight. These overall trends disguised highly variable individual-level BMI trajectories; over one fifth of adults with T2D and almost a quarter of those with hypertension gained weight rapidly (>0.5kg/m^2^/year). The characteristics associated with rapid weight gain were similar in both populations, with the highest likelihood amongst women, younger adults, those living in the most deprived areas, adults of white ethnicity and those living with comorbid mental health conditions.

#### Research in context

Data on weight trajectories amongst people living with hypertension are lacking, and we present novel evidence that, on average, this population maintained a stable weight during the pandemic. Our finding that, on average, adults living with T2D in England lost weight during the pandemic has several possible disease-specific explanatory pathways. Diabetes and obesity were identified as independent risk factors for severe disease with COVID-19[6]. Therefore, people with T2D may have made positive behavioural changes promoting weight loss, as seen in other European settings[14, 15]. We found non-insulin treatment reduced the likelihood of rapid weight gain. Notably, prescription of novel diabetes drugs associated with weight loss (including sodium glucose co-transporter-2 inhibitors) increased in England during the pandemic (Appendix Supplementary 4). Alternatively, weight loss can be a sign of poorly controlled diabetes and may reflect worsening glycaemic control amongst adults with T2D. We would welcome further research investigating the impact of the pandemic on glycaemic control.

A high proportion of adults with T2D and/or hypertension gained weight rapidly, with the likelihood of rapid pandemic weight gain greatest amongst women, young adults, those living in more deprived areas and those with mental health conditions. These patterns mirror those seen in the general population[9] and provide further evidence of pandemic health inequalities, which have been described in relation to sex, ethnicity, wealth, employment and education[16]. Our results support previous reports that young adults are at high risk of weight gain[8, 9]. The increased odds amongst women may reflect gender disparities in patterns of employment and caring responsibilities during the pandemic[17]. Associations with deprivation are likely multifactorial in aetiology, including food poverty, reduced opportunity for physical activity and an increased burden of physical and mental health conditions[18]. Our novel findings that comorbid mental health conditions increased odds of rapid weight gain amongst adults with T2D and/or hypertension may reflect associations between disordered eating, reduced physical activity and poor mental health exacerbated by the pandemic[19, 20].

#### Strength and limitations

The key strengths of this study are the quality, scale and representativeness of the data used[21]. In England, primary care hosts and records data from over 90% of all patient consultations in the National Health Service[22]. OpenSAFELY-TPP provides access to primary care records of roughly 40% of the population[21]. The large sample size supports the generalisability of these findings and allowed us to investigate patterns of weight gain in many sociodemographic and clinical strata, which uncovered novel findings.

Although BMI data are not entered into the EHR following a predefined study protocol, routine EHRs provide BMI trajectory estimates comparable to those from population-based surveys[23]. Adults with T2D are more likely to have their BMI recorded as this is part of the annual diabetes review[24].

Restructuring of healthcare services during the pandemic may have introduced new bias[9, 25]. However, we report a high proportion of adults with T2D continued to have BMI recorded. (Table Supplementary 1) The BMI data for adults living with hypertension were less complete and may be at greater risk of information bias. (Table Supplementary 2).

Our work has limitations. Increased BMI self-reporting during remote consultations may have introduced biases[26]. We conducted an analysis of patients who were registered with a general practitioner at the time of data extraction. This provides robust estimates of the current burden of obesity and rapid weight gain but may have introduced survival bias. We undertook a complete case analysis, which may preclude the generalisability of these findings to individuals with missing ethnicity or IMD data. Notably, a low proportion of adults had missing data in these fields.

#### Policy Implications

We have identified clear characteristics associated with odds of rapid weight gain which could enable targeting of existing weight management interventions[10]. General practitioners are at the front line of healthcare in the UK. They play a key role in supporting weight management through brief interventions and onward referral to weight management programmes [27]. We highlight the population subgroups that were at greatest risk of unhealthy patterns of weight gain during the pandemic and may consequently benefit the most from weight management interventions. Our findings, that comorbid mental health conditions are both prevalent amongst adults living with T2D and/or hypertension and are associated with increased odds of rapid weight gain, highlight the complex interplay between physical health, mental health and weight. Future research and policy in this field must incorporate sociodemographic, physical and mental health characteristics in prioritisation and implementation of interventions to ensure equity.

## Data Availability

Detailed pseudonymised patient data is potentially re-identifiable and therefore not shared. All code for data management and analysis, as well as codelists, are shared openly for review and re-use under MIT open license, available at https://github.com/opensafely/BMI-and-Metabolic-Markers.

## Ethical Approval

The study was approved by the London School of Hygiene & Tropical Medicine Ethics Board (reference 26536).

## Data Sharing

Access to the underlying identifiable and potentially re-identifiable pseudonymised electronic health record data is tightly governed by various legislative and regulatory frameworks, and restricted by best practice. The data in OpenSAFELY-TPP is drawn from General Practice data across England where TPP is the data processor.

TPP developers (Chris Bates, Jonathan Cockburn, John Parry, Frank Hester, and Sam Harper) initiate an automated process to create pseudonymised records in the core OpenSAFELY database, which are copies of key structured data tables in the identifiable records. These pseudonymised records are linked onto key external data resources that have also been pseudonymised via SHA-512 one-way hashing of NHS numbers using a shared salt. Bennett Institute for Applied Data Science developers and PIs (Ben Goldacre, Liam Smeeth, Caroline E Morton, Seb Bacon, Alex J Walker, William Hulme, Helen J Curtis, David Evans, Peter Inglesby, Simon Davy, George Hickman, Krishnan Bhaskaran, and Christopher T Rentsch) holding contracts with NHS England have access to the OpenSAFELY pseudonymised data tables as needed to develop the OpenSAFELY tools.

These tools in turn enable researchers with OpenSAFELY Data Access Agreements to write and execute code for data management and data analysis without direct access to the underlying raw pseudonymised patient data, and to review the outputs of this code. All code for the full data management pipeline—from raw data to completed results for this analysis—and for the OpenSAFELY platform as a whole is available for review at https://github.com/OpenSAFELY. The data management and analysis code for this paper was led by MS and RYP and is available for scientific review and re-use under MIT open licence. https://github.com/opensafely/BMI-and-Metabolic-Markers.

## Acknowledgements

We are very grateful for all the support received from the TPP Technical Operations team throughout this work, and for generous assistance from the information governance and database teams at NHS England and the NHS England Transformation Directorate. KK is supported by the National Institute for Health Research (NIHR) Applied Research Collaboration East Midlands (ARC EM), NIHR Global Research Centre for Multiple Long Term Conditions and the NIHR Leicester Biomedical Research Centre (BRC).

## Authors Contribution Statement

All authors contributed to this manuscript. BG contributed to the funding acquisition for the analysis platform. MS, RYP, RM, CEM, BMK, KK, JV and SF contributed to the conceptualisation of the study. MS, RYP, CEM, SB, AM, BG, JM, ID, PI, WJH, RR, BMK and SF were involved in project administration underpinning this analysis. CEM, SB, AM, JM, ID, PI, WJH, and BMK contributed to the development and maintenance of the OpenSAFELY-TPP platform. MS, RYP, CEM, SB, AM, BG, ID, PI and WJH were involved in curation of the data used in the analysis. MS, RYP, SVE, FE, DS, RM and SF contributed to the development and design of the statistical methodology used in these analyses. MS conducted the formal analysis including the application of statistical techniques with technical support from RYP and further support and supervision from DS, SVE, RM and SF. RYP, SVE, WJH, KK, RM, JV, BMK and SF contributed to supervision of different parts of the analysis including support to use the platform, support in the analyses and interpretation of the study findings. MS wrote the original draft. All authors contributed to further review and editing of the draft.

