## Supplementary for "Weight trends amongst adults with diabetes or hypertension during the COVID-19 pandemic: an observational study using OpenSAFELY"

*Figure S1. Adjusted odds of rapid weight gain amongst adults living with type 2 diabetes in England after the onset of the COVID-19 pandemic adjusted for age, sex, gender, IMD, ethnicity and diabetes medication.* 5

Appendix S1. Information Governance Statement

NHS England is the data controller of the NHS England OpenSAFELY COVID-19 Service; TPP is the data processor; all study authors using OpenSAFELY have the approval of NHS England.[1] This implementation of OpenSAFELY is hosted within the TPP environment which is accredited to the ISO 27001 information security standard and is NHS IG Toolkit compliant;[2] Patient data has been pseudonymised for analysis and linkage using industry standard cryptographic hashing techniques; all pseudonymised datasets transmitted for linkage onto OpenSAFELY are encrypted; access to the NHS England OpenSAFELY COVID-19 service is via a virtual private network (VPN) connection; the researchers hold contracts with NHS England and only access the platform to initiate database queries and statistical models; all database activity is logged; only aggregate statistical outputs leave the platform environment following best practice for anonymisation of results such as statistical disclosure control for low cell counts.[3] The service adheres to the obligations of the UK General Data Protection Regulation (UK GDPR) and the Data Protection Act 2018. The service previously operated under notices initially issued in February 2020 by the the Secretary of State under Regulation 3(4) of the Health Service (Control of Patient Information) Regulations 2002 (COPI Regulations), which required organisations to process confidential patient information for COVID-19 purposes; this set aside the requirement for patient consent.[4] As of 1 July 2023, the Secretary of State has requested that NHS England continue to operate the Service under the COVID-19 Directions 2020.[5] In some cases of data sharing, the common law duty of confidence is met using, for example, patient consent or support from the Health Research Authority Confidentiality Advisory Group.[6] Taken together, these provide the legal bases to link patient datasets using the service. GP practices, which provide access to the primary care data, are required to share relevant health information to support the public health response to the pandemic, and have been informed of how the service operates. This study was supported by Dr Jonathan Valabhji, clinical director for diabetes and obesity at NHS England, as senior sponsor. The study was approved by the London School of Hygiene & Tropical Medicine Ethics Board (reference 26536)

Appendix S2. Calculation of rate of weight gain

BMIs were classified into time periods: period-1 (March 2018 - February 2020); and period-2 (March 2020 - February 2022). For patients with more than one BMI measure from each time period, a random BMI measure was selected from each time period. Where data were available, BMI data from period 1 and period 2 were used to calculate the rate of BMI change/year (δ) assuming a linear trend (Katsoulis, Lai, et al. 2021).

The rate of weight gain before the onset of the pandemic (δ-prepandemic) was calculated using data from period-0 (March 2015 - February 2018) and period-1. (Supplementary Figure 1). The findings of the prepandemic analysis are presented in Supplementary Figure 8.

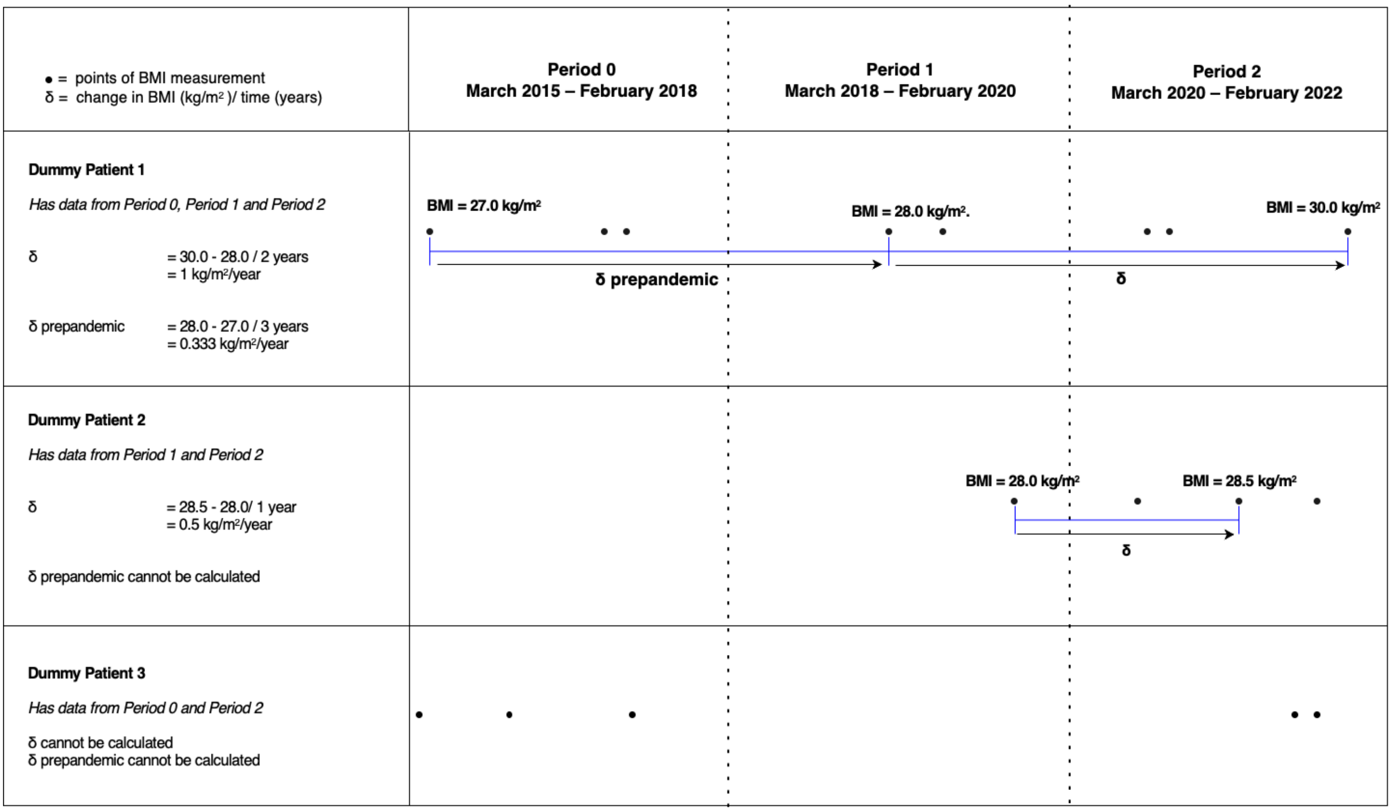

### Appendix S3. Covariates

Covariates were chosen by clinical consensus, guided by data availability and known potential predictors of BMI change. Covariates included age (18-29 years, 30-39 years, 40-49 years, 50-59 years, 60-69 years, 70-79 years, 80 ≤ 90 years), sex (female or male). We used ethnicity categories based on the 2001 UK Census definitions which were further collapsed into the following subgroups to reduce risk of disclosure from small groups (White (White British, White Irish, Other White), Black (Black African, Black Caribbean, Other Black), South Asian (Indian, Pakistani, Bangladeshi), and Chinese/Other (Chinese, Other Asian, Mixed White/Black African, Mixed White/Black Caribbean, Mixed White/Asian, Other). We measured deprivation using the most recent patient postcode-derived Index of Multiple Deprivation (IMD; by quintiles from those living in the most deprived 20% of households to the least deprived 20%). We classified whether individuals had comorbidities, based on the presence or absence of codes for the following long term conditions ever-recorded: hypertension, type 1 diabetes, type 2 diabetes, asthma, chronic obstructive pulmonary disease, anxiety and depression, serious mental illness (psychosis or bipolar disorder), learning difficulties, dementia, cardiovascular disease, and stroke and transient ischaemic attack. Adults with type 2 diabetes were further classified based on their prescribed diabetes medication regimen into the following three groups: insulin containing regimens (insulin), regimens that did not include insulin (non-insulin), and those who were not prescribed pharmacological treatment.

Figure S1. Adjusted odds of rapid weight gain amongst adults living with type 2 diabetes in England after the onset of the COVID-19 pandemic adjusted for age, sex, gender, IMD, ethnicity and diabetes medication

| Population Subgroup | N (%) | Prevalence (%) | aOR (95% CI) |
|---|---|---|---|
| All | 793,770 (100.0) | 20.7 | |
| Sex | | | |
| Female | 345,050 (43.5) | 23.0 | 1 |
| Male | 448,725 (56.5) | 19.0 | 0.80 (0.79 to 0.81) |
| Age in years | | | |
| 18-29 | 3,310 (0.4) | 30.4 | 1 |
| 30-39 | 19,790 (2.5) | 25.5 | 0.84 (0.78 to 0.92) |
| 40-49 | 68,410 (8.6) | 22.6 | 0.76 (0.70 to 0.82) |
| 50-59 | 163,050 (20.5) | 22.2 | 0.72 (0.67 to 0.78) |
| 60-69 | 218,335 (27.5) | 21.3 | 0.67 (0.62 to 0.72) |
| 70-79 | 214,905 (27.1) | 19.5 | 0.58 (0.53 to 0.62) |
| 80-90 | 105,970 (13.4) | 17.4 | 0.48 (0.44 to 0.52) |
| Ethnicity | | | |
| White | 640,190 (80.7) | 21.3 | 1 |
| Black | 23,935 (3.0) | 22.2 | 0.95 (0.92 to 0.98) |
| South Asian | 89,750 (11.3) | 17.3 | 0.72 (0.71 to 0.73) |
| Mixed | 7,715 (1.0) | 22.6 | 0.99 (0.94 to 1.05) |
| Chinese and other | 32,180 (4.1) | 17.8 | 0.76 (0.74 to 0.78) |
| Index of Multiple Deprivation | | | |
| 1 (most deprived) | 191,770 (24.2) | 22.0 | 1 |
| 2 | 174,110 (21.9) | 21.1 | 0.96 (0.95 to 0.98) |
| 3 | 168,865 (21.3) | 20.6 | 0.93 (0.92 to 0.95) |
| 4 | 142,585 (18.0) | 20.0 | 0.90 (0.88 to 0.92) |
| 5 (least deprived) | 116,445 (14.7) | 19.3 | 0.86 (0.85 to 0.88) |
| Region | | | |
| East of England | 164,595 (20.7) | 20.7 | 1 |
| East Midlands | 142,570 (18.0) | 20.5 | 0.98 (0.97 to 1.00) |
| London | 57,325 (7.2) | 19.3 | 0.99 (0.97 to 1.02) |
| North East | 40,460 (5.1) | 20.5 | 0.95 (0.93 to 0.98) |
| North West | 77,985 (9.8) | 22.0 | 1.01 (0.99 to 1.03) |
| South East | 45,895 (5.8) | 20.6 | 0.97 (0.95 to 1.00) |
| South West | 104,350 (13.1) | 21.5 | 1.03 (1.01 to 1.05) |
| West Midlands | 38,690 (4.9) | 20.4 | 0.97 (0.94 to 1.00) |
| Yorkshire and The Humber | 121,900 (15.4) | 20.6 | 0.98 (0.96 to 0.99) |
| Long Term Condition | | | |
| Hypertension | 520,365 (65.6) | 20.7 | 1.06 (1.05 to 1.07) |
| Chronic Cardiac Disease | 181,965 (22.9) | 20.6 | 1.06 (1.04 to 1.07) |
| Learning Disability | 7,030 (0.9) | 25.4 | 1.13 (1.07 to 1.19) |
| Depression | 208,775 (26.3) | 23.5 | 1.11 (1.10 to 1.13) |
| Dementia | 17,345 (2.2) | 21.4 | 1.15 (1.11 to 1.20) |
| Serious Mental Illness | 20,065 (2.5) | 25.7 | 1.22 (1.18 to 1.26) |
| Asthma | 155,055 (19.5) | 22.4 | 1.06 (1.04 to 1.07) |
| COPD | 66,110 (8.3) | 22.2 | 1.11 (1.09 to 1.13) |
| Stroke and TIA | 64,300 (8.1) | 20.7 | 1.04 (1.02 to 1.07) |
| Diabetic Medication | | | |
| No medication | 193,125 (24.3) | 24.0 | 1 |
| Non-insulin medication | 494,220 (62.3) | 18.0 | 0.68 (0.67 to 0.69) |
| Insulin | 106,425 (13.4) | 27.4 | 1.16 (1.14 to 1.18) |

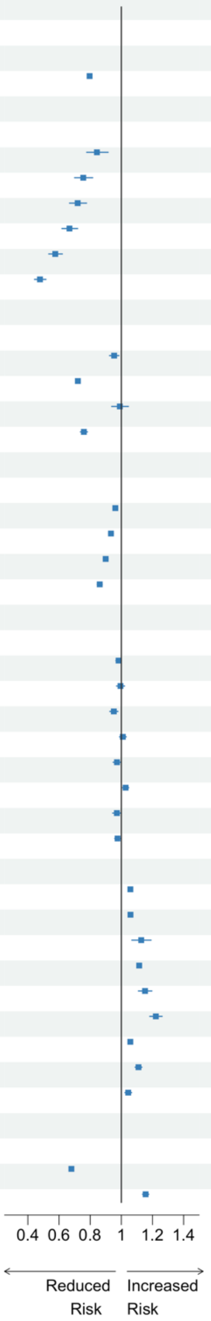

Rapid weight gain: > 0.5 kilograms/metre$^2$/year. N (%): Total number (N) and percentage (%) from each population strata contributing to the analysis. Prevalence (%): percentage who gained weight rapidly. aOR: adjusted Odds Ratio generated in logistic regression models adjusted for age, sex, ethnicity, Index of Multiple Deprivation and diabetic medication. 95% CI: 95% Confidence Interval of aOR. COPD: Chronic Obstructive Pulmonary Disease. TIA: Transient Ischaemic Attack.

Appendix 4. Population level diabetes medication prescription trends

The relative prescription of novel antidiabetic drugs, including sodium glucose co-transporter-2 (SGLT-2) inhibitors and glucagon-like peptide-1 (GLP-1) receptor agonists which are associated with weight loss, in England since the onset of the pandemic. The following graphs have been created using [https://openprescribing.net/](https://openprescribing.net/) and demonstrate the frequency of prescription of of SGLT-2 inhibitors and GLP-1 receptor agonists per 1000 total antidiabetic prescriptions, compared to more established antidiabetic drugs such as metformin and gliclazide. Although the total number of prescriptions of novel agents are less than the more established drugs, there is a trend towards increased prescription of novel agents and decreased prescription of metformin and gliclazide over time.

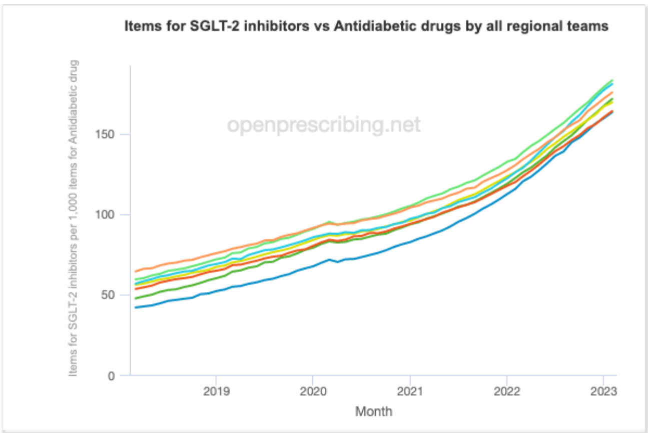

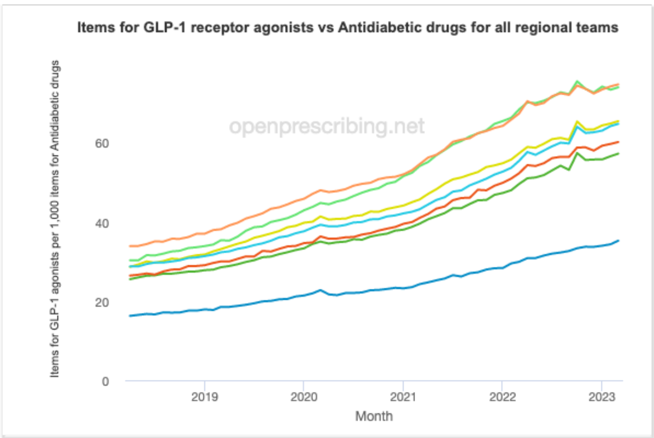

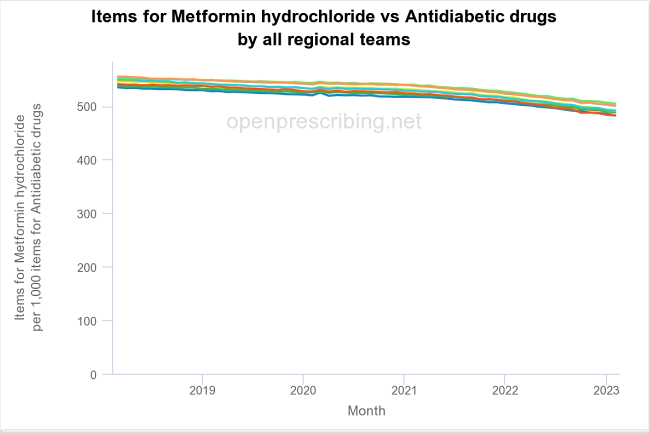

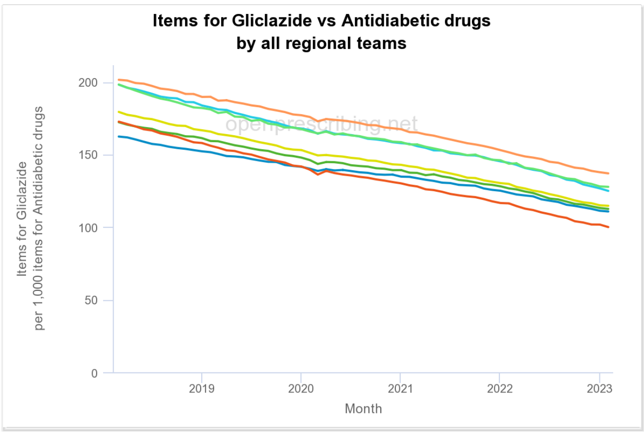

Table S1. Demographics of adults with type 2 diabetes contributing data to each stage of the analysis

| | Total Population | Cross Sectional BMI Analysis | | Rate of weight gain analysis | |
|---|---|---|---|---|---|
| | N (%) | N (%) | BMI data (%) | N (%) | BMI data (%) |
| **All** | 1,231,455 | 1,164,805 | 94.6 | 793,770 | 64.5 |
| **Sex** | | | | | |
| Female | 540,925 (43.9) | 510,690 (43.7) | 94.4 | 345,050 (43.5) | 63.8 |
| Male | 690,530 (56.1) | 654,115 (56.3) | 94.7 | 448,725 (56.5) | 65.0 |
| **Age in years** | | | | | |
| 18-29 | 6,445 (0.5) | 5,325 (0.3) | 82.6 | 3,310 (0.4) | 51.4 |
| 30-39 | 32,970 (2.7) | 29,030 (2.0) | 88.0 | 19,790 (2.5) | 60.0 |
| 40-49 | 105,340 (8.6) | 96,435 (7.3) | 91.5 | 68,410 (8.6) | 64.9 |
| 50-59 | 243,280 (19.8) | 227,470 (18.6) | 93.5 | 163,050 (20.5) | 67.0 |
| 60-69 | 325,060 (26.4) | 309,685 (26.8) | 95.3 | 218,335 (27.5) | 67.2 |
| 70-79 | 334,850 (27.2) | 322,085 (28.9) | 96.2 | 214,905 (27.1) | 64.2 |
| 80-90 | 183,510 (14.9) | 174,780 (16.1) | 95.2 | 105,970 (13.4) | 57.7 |
| **Ethnicity** | | | | | |
| White British | 902,010 (76.0) | 857,790 (76.8) | 95.1 | | |
| White Irish | 6,225 (0.5) | 5,910 (0.5) | 94.9 | | |
| Other White | 56,275 (4.7) | 51,935 (4.5) | 92.3 | | |
| Black Caribbean | 13,400 (1.1) | 12,675 (1.1) | 94.6 | | |
| Black African | 16,315 (1.4) | 14,900 (1.2) | 91.3 | | |
| Other Black | 6,835 (0.6) | 6,340 (0.5) | 92.8 | | |
| Indian | 65,435 (5.5) | 61,690 (5.4) | 94.3 | | |
| Pakistani | 48,950 (4.1) | 46,385 (4.1) | 94.8 | | |
| Bangladeshi | 12,975 (1.1) | 12,290 (1.1) | 94.7 | | |
| Chinese | 4,245 (0.4) | 3,940 (0.3) | 92.8 | | |
| Other Asian | 26,595 (2.2) | 25,035 (2.1) | 94.1 | | |
| Mixed White/Black Caribbean | 3,015 (0.3) | 2,830 (0.2) | 93.9 | | |
| Mixed White/Black African | 2,325 (0.2) | 2,140 (0.2) | 92.0 | | |
| Mixed White/Asian | 2,440 (0.2) | 2,290 (0.2) | 93.9 | | |
| Other Mixed | 3,920 (0.3) | 3,635 (0.3) | 92.7 | | |
| Other | 16,170 (1.4) | 14,895 (1.2) | 92.1 | | |
| NA | 44,330 (3.6) | 40,135 (3.4) | 90.5 | | |
| **Collapsed Ethnicity** | | | | | |
| White | 964,510 (81.2) | 915,635 (81.8) | 94.9 | 640,190 (80.7) | 66.4 |
| Black | 36,550 (3.1) | 33,915 (2.9) | 92.8 | 23,935 (3.0) | 65.5 |
| South Asian | 127,360 (10.7) | 120,365 (10.6) | 94.5 | 89,750 (11.3) | 70.5 |
| Mixed | 11,700 (1.0) | 10,895 (0.9) | 93.1 | 7,715 (1.0) | 65.9 |
| Chinese/Other | 47,010 (4.0) | 43,870 (3.7) | 93.3 | 32,180 (4.1) | 68.5 |
| **Index of Multiple Deprivation** | | | | | |
| 1 (most deprived) | 285,035 (23.6) | 268,565 (23.5) | 94.2 | 191,770 (24.2) | 67.3 |
| 2 | 263,375 (21.8) | 248,485 (21.7) | 94.3 | 174,110 (21.9) | 66.1 |
| 3 | 256,550 (21.3) | 243,505 (21.3) | 94.9 | 168,865 (21.3) | 65.8 |
| 4 | 220,190 (18.2) | 208,770 (18.3) | 94.8 | 142,585 (18.0) | 64.8 |
| 5 (least deprived) | 181,945 (15.1) | 172,990 (15.2) | 95.1 | 116,445 (14.7) | 64.0 |
| NA | 24,360 (2.0) | 22,485 (1.7) | 92.3 | | |
| **Region** | | | | | |
| East of England | 266,540 (21.6) | 250,010 (21.4) | 93.8 | 164,595 (20.7) | 61.8 |
| East Midlands | 225,050 (18.3) | 211,255 (18.3) | 93.9 | 142,570 (18.0) | 63.4 |
| London | 79,705 (6.5) | 76,270 (6.3) | 95.7 | 57,325 (7.2) | 71.9 |
| North East | 60,095 (4.9) | 57,040 (5.0) | 94.9 | 40,460 (5.1) | 67.3 |
| North West | 117,290 (9.5) | 111,485 (9.6) | 95.1 | 77,985 (9.8) | 66.5 |
| South East | 75,645 (6.1) | 71,155 (6.0) | 94.1 | 45,895 (5.8) | 60.7 |
| South West | 160,600 (13.0) | 153,195 (13.0) | 95.4 | 104,350 (13.1) | 65.0 |
| West Midlands | 59,825 (4.9) | 56,350 (4.9) | 94.2 | 38,690 (4.9) | 64.7 |
| Yorkshire and The Humber | 186,705 (15.2) | 178,035 (15.5) | 95.4 | 121,900 (15.4) | 65.3 |
| **Long Term Condition** | | | | | |
| Hypertension | 792,630 (64.4) | 753,690 (64.4) | 95.1 | 520,365 (65.6) | 65.7 |
| Chronic Cardiac Disease | 275,510 (22.4) | 259,710 (21.1) | 94.3 | 181,965 (22.9) | 66.0 |
| Learning Disability | 9,290 (0.8) | 8,910 (0.7) | 95.9 | 7,030 (0.9) | 75.7 |
| Depression | 316,555 (25.7) | 297,815 (24.7) | 94.1 | 208,775 (26.3) | 66.0 |
| Dementia | 31,790 (2.6) | 26,830 (1.8) | 84.4 | 17,345 (2.2) | 54.6 |
| Serious Mental Illness | 27,555 (2.2) | 26,360 (2.1) | 95.7 | 20,065 (2.5) | 72.8 |
| Asthma | 230,390 (18.7) | 218,300 (18.2) | 94.8 | 155,055 (19.5) | 67.3 |
| COPD | 100,305 (8.1) | 94,745 (7.5) | 94.5 | 66,110 (8.3) | 65.9 |
| Stroke and TIA | 103,420 (8.4) | 94,635 (7.4) | 91.5 | 64,300 (8.1) | 62.2 |

BMI: Body Mass Index. N(%): refers to the total number (N) and percentage (%) of individuals contributing to the analysis from each population subgroup. BMI data (%): refers to the percentage of individuals in each population subgroup contributing data to the analysis. TIA: Transient Ischaemic Attack. Serious Mental Illness: Includes Bipolar Disorder and Psychosis.

Table S2: Demographics of adults living with hypertension contributing data to each stage of the analysis

| | Total Population | Cross Sectional BMI Analysis | | Rate of weight gain Analysis | |
|---|---|---|---|---|---|
| | N(%) | N(%) | BMI data available (%) | N(%) | BMI data available (%) |
| **All** | 3,558,405 (100.0) | 2,868,175 (100.0) | 80.6 | 1,436,070 (100.0) | 40.4 |
| Sex | | | | | |
| Female | 1,767,845 (49.7) | 1,428,840 (50.1) | 80.8 | 700,230 (48.8) | 39.6 |
| Male | 1,790,560 (50.3) | 1,439,330 (49.9) | 80.4 | 735,840 (51.2) | 41.1 |
| **Age in years** | | | | | |
| 18-29 | 17,765 (0.5) | 10,560 (0.3) | 59.4 | 4,725 (0.3) | 26.6 |
| 30-39 | 74,180 (2.1) | 48,295 (1.4) | 65.1 | 23,230 (1.6) | 31.3 |
| 40-49 | 249,180 (7.0) | 176,625 (5.5) | 70.9 | 89,400 (6.2) | 35.9 |
| 50-59 | 649,915 (18.3) | 494,130 (16.4) | 76.0 | 253,110 (17.6) | 38.9 |
| 60-69 | 894,390 (25.1) | 727,430 (25.3) | 81.3 | 378,515 (26.4) | 42.3 |
| 70-79 | 1,057,325 (29.7) | 891,030 (32.1) | 84.3 | 446,120 (31.1) | 42.2 |
| 80-90 | 615,645 (17.3) | 520,120 (19.2) | 84.5 | 240,965 (16.8) | 39.1 |
| **Ethnicity** | | | | | |
| White British | 2,858,145 (84.3) | 2,341,100 (85.3) | 81.9 | | |
| White Irish | 22,675 (0.7) | 18,060 (0.7) | 79.6 | | |
| Other White | 179,285 (5.3) | 133,125 (4.7) | 74.3 | | |
| Black Caribbean | 28,065 (0.8) | 23,355 (0.9) | 83.2 | | |
| Black African | 35,335 (1.0) | 26,150 (0.9) | 74.0 | | |
| Other Black | 14,905 (0.4) | 11,385 (0.4) | 76.4 | | |
| Indian | 90,925 (2.7) | 76,020 (2.7) | 83.6 | | |
| Pakistani | 49,395 (1.5) | 43,120 (1.5) | 87.3 | | |
| Bangladeshi | 11,625 (0.3) | 9,930 (0.3) | 85.4 | | |
| Chinese | 8,510 (0.3) | 6,355 (0.2) | 74.7 | | |
| Other Asian | 37,150 (1.1) | 30,075 (1.0) | 81.0 | | |
| Mixed White/Black Caribbean | 6,805 (0.2) | 5,470 (0.2) | 80.4 | | |
| Mixed White/Black African | 5,445 (0.2) | 4,025 (0.1) | 73.9 | | |
| Mixed White/Asian | 4,045 (0.1) | 3,175 (0.1) | 78.5 | | |
| Other Mixed | 8,175 (0.2) | 6,240 (0.2) | 76.3 | | |
| Other | 30,680 (0.9) | 23,680 (0.8) | 77.2 | | |
| NA | 167,250 (4.7) | 106,910 (3.8) | 63.9 | | |
| **Collapsed Ethnic Groups** | | | | | |
| White | 3,060,105 (90.2) | 2,492,285 (90.6) | 81.4 | 1,277,245 (88.9) | 41.7 |
| Black | 78,305 (2.3) | 60,890 (2.1) | 77.8 | 32,415 (2.3) | 41.4 |
| South Asian | 151,945 (4.5) | 129,070 (4.5) | 84.9 | 81,670 (5.7) | 53.7 |
| Mixed | 24,470 (0.7) | 18,910 (0.7) | 77.3 | 10,040 (0.7) | 41.0 |
| Chinese/Other | 76,340 (2.3) | 60,110 (2.1) | 78.7 | 34,695 (2.4) | 45.4 |
| **Index of Multiple Deprivation** | | | | | |
| 1 (most deprived) | 619,365 (17.7) | 518,465 (18.4) | 83.7 | 290,830 (20.3) | 47.0 |
| 2 | 675,975 (19.4) | 551,080 (19.6) | 81.5 | 290,285 (20.2) | 42.9 |
| 3 | 770,590 (22.1) | 622,255 (22.0) | 80.8 | 315,530 (22.0) | 40.9 |
| 4 | 741,380 (21.2) | 589,130 (20.9) | 79.5 | 287,720 (20.0) | 38.8 |
| 5 (least deprived) | 682,595 (19.6) | 534,430 (19.0) | 78.3 | 251,700 (17.5) | 36.9 |
| NA | 68,495 (1.9) | 52,815 (1.6) | 77.1 | NA (NA) | NA |
| **Region** | | | | | |
| East of England | 813,655 (22.9) | 615,870 (21.7) | 75.7 | 275,225 (19.2) | 33.8 |
| East Midlands | 632,290 (17.8) | 516,870 (18.2) | 81.7 | 265,845 (18.5) | 42.0 |
| London | 181,005 (5.1) | 142,775 (4.9) | 78.9 | 79,970 (5.6) | 44.2 |
| North East | 175,945 (4.9) | 147,765 (5.2) | 84.0 | 80,965 (5.6) | 46.0 |
| North West | 347,655 (9.8) | 302,970 (10.5) | 87.1 | 173,545 (12.1) | 49.9 |
| South East | 231,260 (6.5) | 172,525 (5.9) | 74.6 | 71,885 (5.0) | 31.1 |
| South West | 519,315 (14.6) | 419,135 (14.3) | 80.7 | 204,570 (14.2) | 39.4 |
| West Midlands | 142,520 (4.0) | 111,650 (3.9) | 78.3 | 54,390 (3.8) | 38.2 |
| Yorkshire and The Humber | 514,760 (14.5) | 438,610 (15.3) | 85.2 | 229,680 (16.0) | 44.6 |
| **Long Term Condition** | | | | | |
| Type 1 Diabetes | 32,690 (0.9) | 30,400 (1.0) | 93.0 | 20,115 (1.4) | 61.5 |
| Type 2 Diabetes | 792,630 (22.3) | 753,690 (24.8) | 95.1 | 520,365 (36.2) | 65.7 |
| Chronic Cardiac Disease | 660,170 (18.6) | 571,865 (18.7) | 86.6 | 329,730 (23.0) | 49.9 |
| Learning Disability | 14,300 (0.4) | 13,385 (0.4) | 93.6 | 9,865 (0.7) | 69.0 |
| Depression | 825,640 (23.2) | 678,795 (22.9) | 82.2 | 359,425 (25.0) | 43.5 |
| Dementia | 78,715 (2.2) | 58,295 (1.4) | 74.1 | 31,490 (2.2) | 40.0 |
| Serious Mental Illness | 45,295 (1.3) | 41,900 (1.3) | 92.5 | 28,420 (2.0) | 62.7 |
| Asthma | 603,260 (17.0) | 512,930 (17.5) | 85.0 | 279,720 (19.5) | 46.4 |
| COPD | 256,525 (7.2) | 228,825 (7.5) | 89.2 | 130,170 (9.1) | 50.7 |
| Stroke and TIA | 297,180 (8.4) | 244,170 (7.7) | 82.2 | 133,430 (9.3) | 44.9 |

BMI: Body Mass Index. N(%): refers to total number (N) and percentage (%) of individuals contributing to the analysis from each population subgroup. BMI data (%): refers to the percentage of individuals in each population subgroup contributing data to the analysis. COPD: Chronic Obstructive Pulmonary Disease. TIA: Transient Ischaemic Attack. Serious Mental Illness: Includes Bipolar Disorder and Psychosis

Table S3: The median Body Mass Index (BMI), and proportion obese, of adults living in England with type 2 diabetes or hypertension during the COVID-19 pandemic

| | Type 2 Diabetes BMI | | | Hypertension BMI | | |
|---|---|---|---|---|---|---|
| | N (%) | Median BMI (IQR) | Obese (%) | N (%) | Median BMI (IQR) | Obese (%) |
| All | 1,164,805 | 30.3 (26.6, 34.8) | 55.5 | 2,868,175 | 29.0 (25.5, 33.3) | 45.2 |
| **Sex** | | | | | | |
| Female | 510,690 (43.8) | 31.1 (26.9, 36.2) | 60.3 | 1,428,840 (49.8) | 29.0 (25.1, 34.0) | 46.3 |
| Male | 654,115 (56.2) | 29.7 (26.5, 33.8) | 51.8 | 1,439,330 (50.2) | 29.0 (25.9, 32.7) | 44.2 |
| **Age in years** | | | | | | |
| 18-29 | 5,325 (0.5) | 35.2 (29.7, 41.1) | 78.3 | 10,560 (0.4) | 29.7 (24.4, 36.5) | 50.6 |
| 30-39 | 29,030 (2.5) | 33.5 (28.6, 39.4) | 74.5 | 48,295 (1.7) | 31.4 (26.7, 37.4) | 61.2 |
| 40-49 | 96,435 (8.3) | 31.9 (27.7, 37.2) | 68.9 | 176,625 (6.2) | 31.3 (27.3, 36.5) | 62.8 |
| 50-59 | 227,470 (19.5) | 31.8 (27.9, 36.7) | 66.4 | 494,130 (17.2) | 30.9 (27.2, 35.6) | 59.3 |
| 60-69 | 309,685 (26.6) | 30.9 (27.3, 35.3) | 60.0 | 727,430 (25.4) | 29.8 (26.3, 34.1) | 51.0 |
| 70-79 | 322,085 (27.7) | 29.6 (26.3, 33.6) | 49.3 | 891,030 (31.1) | 28.4 (25.2, 32.2) | 39.3 |
| 80-90 | 174,780 (15.0) | 27.6 (24.6, 31.1) | 33.7 | 520,120 (18.1) | 26.6 (23.7, 30.0) | 26.4 |
| Ethnicity | | | | | | |
| White British | 857,790 (76.3) | 30.8 (27.1, 35.3) | 55.6 | 2,341,100 (84.8) | 29.0 (25.6, 33.4) | 43.8 |
| White Irish | 5,910 (0.5) | 29.8 (26.4, 33.9) | 48.8 | 18,060 (0.7) | 28.4 (25.0, 32.3) | 39.0 |
| Other White | 51,935 (4.6) | 30.9 (27.3, 35.4) | 56.9 | 133,125 (4.8) | 29.4 (25.9, 33.8) | 46.7 |
| Black Caribbean | 12,675 (1.1) | 29.8 (26.3, 34.4) | 67.0 | 23,355 (0.8) | 29.5 (26.0, 34.1) | 64.7 |
| Black African | 14,900 (1.3) | 29.7 (26.3, 33.7) | 66.7 | 26,150 (0.9) | 30.1 (26.9, 34.3) | 71.1 |
| Other Black | 6,340 (0.6) | 29.9 (26.4, 34.4) | 67.7 | 11,385 (0.4) | 29.9 (26.4, 34.3) | 67.9 |
| Indian | 61,690 (5.5) | 27.4 (24.6, 31.0) | 49.7 | 76,020 (2.8) | 27.5 (24.6, 31.0) | 50.1 |
| Pakistani | 46,385 (4.1) | 28.9 (25.8, 32.9) | 61.5 | 43,120 (1.6) | 29.2 (26.0, 33.2) | 63.5 |
| Bangladeshi | 12,290 (1.1) | 26.6 (24.1, 29.7) | 42.5 | 9,930 (0.4) | 27.1 (24.5, 30.3) | 46.7 |
| Chinese | 3,940 (0.4) | 25.2 (22.8, 28.1) | 29.7 | 6,355 (0.2) | 25.2 (22.7, 28.1) | 29.6 |
| Other Asian | 25,035 (2.2) | 27.5 (24.7, 30.9) | 50.2 | 30,075 (1.1) | 27.6 (24.7, 30.9) | 50.9 |
| Mixed White/Black Caribbean | 2,830 (0.3) | 30.7 (26.8, 35.5) | 71.2 | 5,470 (0.2) | 30.1 (26.1, 34.8) | 66.6 |
| Mixed White/Black African | 2,140 (0.2) | 30.2 (27.0, 34.6) | 71.7 | 4,025 (0.1) | 30.3 (26.7, 34.4) | 70.1 |
| Mixed White/Asian | 2,290 (0.2) | 28.2 (25.1, 32.5) | 56.3 | 3,175 (0.1) | 28.0 (24.9, 32.2) | 54.5 |
| Other Mixed | 3,635 (0.3) | 29.6 (26.0, 34.2) | 64.8 | 6,240 (0.2) | 29.2 (25.6, 33.6) | 61.9 |
| Other | 14,895 (1.3) | 29.0 (25.8, 33.1) | 62.0 | 23,680 (0.9) | 28.7 (25.5, 32.9) | 60.1 |
| NA | 40,135 (3.4) | 30.1 (26.7, 34.4) | 51.2 | 106,910 (3.7) | 29.1 (25.7, 33.2) | 43.8 |
| **IMD** | | | | | | |
| 1 (most deprived) | 268,565 (23.5) | 31.0 (27.1, 35.8) | 61.9 | 518,465 (18.4) | 30.1 (26.2, 34.8) | 53.9 |
| 2 | 248,485 (21.8) | 30.4 (26.7, 35.0) | 57.4 | 551,080 (19.6) | 29.4 (25.8, 33.8) | 49.0 |
| 3 | 243,505 (21.3) | 30.1 (26.6, 34.6) | 54.4 | 622,255 (22.1) | 28.9 (25.5, 33.1) | 44.4 |
| 4 | 208,770 (18.3) | 30.0 (26.5, 34.2) | 52.1 | 589,130 (20.9) | 28.6 (25.3, 32.7) | 41.5 |
| 5 (least deprived) | 172,990 (15.1) | 29.5 (26.1, 33.6) | 48.4 | 534,430 (19.0) | 28.1 (25.0, 32.0) | 37.7 |
| NA | 22,485 (1.9) | 30.5 (26.9, 35.1) | 57.4 | 52,815 (1.8) | 29.2 (25.7, 33.6) | 46.9 |
| **Region** | | | | | | |
| East of England | 250,010 (21.5) | 30.3 (26.7, 34.8) | 54.7 | 615,870 (21.5) | 29.0 (25.5, 33.3) | 44.7 |
| East Midlands | 211,255 (18.1) | 30.4 (26.7, 34.9) | 56.2 | 516,870 (18.0) | 29.1 (25.6, 33.5) | 46.2 |
| London | 76,270 (6.5) | 28.4 (25.2, 32.4) | 51.7 | 142,775 (5.0) | 28.1 (24.9, 32.1) | 46.9 |
| North East | 57,040 (4.9) | 30.9 (27.2, 35.5) | 58.4 | 147,765 (5.2) | 29.5 (25.9, 34.0) | 48.0 |
| North West | 111,485 (9.6) | 31.0 (27.3, 35.6) | 58.4 | 302,970 (10.6) | 29.4 (25.9, 33.7) | 46.8 |
| South East | 71,155 (6.1) | 30.0 (26.4, 34.4) | 52.1 | 172,525 (6.0) | 28.6 (25.2, 32.8) | 41.6 |
| South West | 153,195 (13.2) | 30.3 (26.7, 34.8) | 53.3 | 419,135 (14.6) | 28.6 (25.3, 32.8) | 41.3 |
| West Midlands | 56,350 (4.8) | 30.0 (26.4, 34.5) | 57.5 | 111,650 (3.9) | 29.2 (25.6, 33.6) | 49.2 |
| Yorkshire and The Humber | 178,035 (15.3) | 30.5 (26.8, 35.0) | 57.4 | 438,610 (15.3) | 29.2 (25.7, 33.5) | 46.4 |
| **Long Term Condition** | | | | | | |
| Hypertension | 753,690 (64.7) | 30.6 (26.9, 35.1) | 57.2 | - | - | - |
| Type 1 Diabetes | - | - | - | 30,400 (1.1) | 28.3 (24.9, 32.3) | 39.4 |
| Type 2 Diabetes | - | - | - | 753,690 (26.3) | 30.6 (26.9, 35.1) | 57.2 |
| Chronic Cardiac Disease | 259,710 (22.3) | 29.9 (26.4, 34.2) | 52.6 | 571,865 (19.9) | 28.7 (25.4, 32.8) | 43.1 |
| Learning Disability | 8,910 (0.8) | 31.5 (27.1, 36.9) | 60.6 | 13,385 (0.5) | 30.7 (26.2, 36.0) | 55.2 |
| Depression | 297,815 (25.6) | 31.6 (27.6, 36.5) | 62.7 | 678,795 (23.7) | 30.0 (26.1, 34.8) | 51.3 |
| Dementia | 26,830 (2.3) | 27.6 (24.1, 31.6) | 36.8 | 58,295 (2.0) | 26.3 (22.8, 30.2) | 27.8 |
| Serious Mental Illness | 26,360 (2.3) | 31.3 (27.2, 36.2) | 62.6 | 41,900 (1.5) | 29.9 (25.9, 34.7) | 52.3 |
| Asthma | 218,300 (18.7) | 31.5 (27.4, 36.5) | 61.9 | 512,930 (17.9) | 29.8 (26.0, 34.6) | 50.6 |
| COPD | 94,745 (8.1) | 30.4 (26.5, 35.0) | 53.9 | 228,825 (8.0) | 28.4 (24.7, 32.9) | 41.1 |
| Stroke and TIA | 94,635 (8.1) | 29.3 (25.8, 33.4) | 47.5 | 244,170 (8.5) | 28.0 (24.7, 31.9) | 37.2 |
| Diabetic Medication | | | | | | |
| No medication | 305,440 (26.2) | 29.7 (26.2, 34.1) | 51.1 | - | - | - |
| Non-insulin medication | 706,670 (60.7) | 30.3 (26.7, 34.8) | 56.1 | - | - | - |
| Insulin | 152,695 (13.1) | 31.3 (27.4, 36.0) | 61.9 | - | - | - |

BMI: Body Mass Index. N(%): refers to total number (N) and percentage (%) of individuals contributing to the analysis from each population subgroup. BMI values are based on most recently BMI recorded in the 5 years preceding March 2022. Obese: Individuals classified as obese based on BMI >=30kg/m$^2$ in White individuals,

>=27.5kg/m$^2$ in black and minority ethnic individuals. COPD: Chronic Obstructive Pulmonary Disease. TIA: Transient Ischaemic Attack. Serious Mental Illness: Includes Bipolar Disorder and Psychosis

Table S4: Unadjusted and adjusted odds of rapid weight gain during the COVID-19 pandemic amongst adults living in England

| | Adults with Type 2 Diabetes | | | | Adults with Hypertension | | | |
|---|---|---|---|---|---|---|---|---|
| | N (%) | Rapid pandemic weight gain % | OR | aOR | N (%) | Rapid pandemic weight gain % | OR | aOR |
| **All** | 793,770 | 20.7 | | | 1,436,070 | 24.7 | | |
| **Sex** | | | | | | | | |
| Female | 345,050(43.5) | 23.0 | 1 | 1 | 700,230(48.8) | 27.2 | 1 | 1 |
| Male | 448,725(56.5) | 19.0 | 0.79(0.78,0.80) | 0.78(0.77,0.79) | 735,840(51.2) | 22.4 | 0.77(0.77,0.78) | 0.75(0.75,0.76) |
| **Age in years** | | | | | | | | |
| 18-29 | 3,310(0.4) | 30.4 | 1 | | 4,725(0.3) | 43.8 | 1 | 1 |
| 30-39 | 19,790(2.5) | 25.5 | 0.79(0.73,0.85) | 0.83(0.77,0.90) | 23,230(1.6) | 39.1 | 0.82(0.77,0.88) | 0.86(0.81,0.92) |
| 40-49 | 68,410(8.6) | 22.6 | 0.67(0.62,0.72) | 0.73(0.68,0.79) | 89,400(6.2) | 33.3 | 0.64(0.60,0.68) | 0.70(0.66,0.74) |
| 50-59 | 163,050(20.5) | 22.2 | 0.66(0.61,0.71) | 0.70(0.65,0.75) | 253,110(17.6) | 30.1 | 0.55(0.52,0.58) | 0.59(0.56,0.63) |
| 60-69 | 218,335(27.5) | 21.3 | 0.62(0.58,0.67) | 0.66(0.61,0.71) | 378,515(26.4) | 25.8 | 0.45(0.42,0.47) | 0.48(0.45,0.51) |
| 70-79 | 214,905(27.1) | 19.5 | 0.56(0.52,0.60) | 0.58(0.54,0.62) | 446,120(31.1) | 21.6 | 0.35(0.33,0.37) | 0.37(0.35,0.39) |
| 80-90 | 105,970(13.4) | 17.4 | 0.48(0.45,0.52) | 0.49(0.46,0.53) | 240,965(16.8) | 18.4 | 0.29(0.27,0.31) | 0.30(0.28,0.31) |
| **Ethnicity** | | | | | | | | |
| White | 640,190(80.7) | 21.3 | 1 | 1 | 1,277,245(88.9) | 25.0 | 1 | 1 |
| Black | 23,935(3.0) | 22.2 | 1.06(1.02,1.09) | 0.95(0.92,0.98) | 32,415(2.3) | 27.2 | 1.12(1.09,1.15) | 0.89(0.87,0.91) |
| South Asian | 89,750(11.3) | 17.3 | 0.78(0.76,0.79) | 0.70(0.69,0.71) | 81,670(5.7) | 20.4 | 0.77(0.75,0.78) | 0.65(0.63,0.66) |
| Mixed | 7,715(1.0) | 22.6 | 1.08(1.03,1.14) | 0.98(0.93,1.04) | 10,040(0.7) | 27.3 | 1.13(1.08,1.18) | 0.92(0.88,0.96) |
| Chinese/ Other | 32,180(4.1) | 17.8 | 0.80(0.78,0.83) | 0.74(0.72,0.76) | 34,695(2.4) | 21.6 | 0.82(0.80,0.85) | 0.70(0.68,0.72) |
| **IMD** | | | | | | | | |
| 1 (most deprived) | 191,770(24.2) | 22.0 | 1 | 1 | 290,830(20.3) | 27.4 | 1 | 1 |
| 2 | 174,110(21.9) | 21.1 | 0.95(0.93,0.96) | 0.96(0.95,0.98) | 290,285(20.2) | 25.7 | 0.92(0.91,0.93) | 0.95(0.94,0.96) |
| 3 | 168,865(21.3) | 20.6 | 0.92(0.91,0.94) | 0.94(0.92,0.95) | 315,530(22.0) | 24.4 | 0.86(0.85,0.87) | 0.90(0.89,0.91) |
| 4 | 142,585(18.0) | 20.0 | 0.89(0.87,0.90) | 0.90(0.89,0.92) | 287,720(20.0) | 23.5 | 0.82(0.81,0.83) | 0.86(0.85,0.87) |
| 5(least deprived) | 116,445(14.7) | 19.3 | 0.85(0.83,0.86) | 0.87(0.85,0.89) | 251,700(17.5) | 22.4 | 0.76(0.76,0.77) | 0.82(0.81,0.83) |
| **Long Term Condition** | | | | | | | | |
| Hypertension | 520,365(65.6) | 20.7 | 0.99(0.98,1.00) | 1.06(1.05,1.08) | | | | |
| Type 2 Diabetes | | | | | 520,365(36.2) | 20.7 | 0.70(0.70,0.71) | 0.74(0.74,0.75) |
| Type 1 Diabetes | | | | | 20,115(1.4) | 28.9 | 1.24(1.20,1.28) | 1.06(1.03,1.09) |
| CCD | 181,965(22.9) | 20.6 | 0.99(0.98,1.00) | 1.09(1.08,1.11) | 329,730(23.0) | 23.0 | 0.88(0.88,0.89) | 1.06(1.05,1.07) |
| LD | 7,030(0.9) | 25.4 | 1.30(1.24,1.38) | 1.13(1.07,1.19) | 9,865(0.7) | 31.9 | 1.43(1.37,1.49) | 1.11(1.06,1.16) |
| Depression | 208,775(26.3) | 23.5 | 1.25(1.23,1.26) | 1.13(1.12,1.15) | 359,425(25.0) | 28.6 | 1.31(1.30,1.32) | 1.15(1.14,1.16) |
| Dementia | 17,345(2.2) | 21.4 | 1.04(1.00,1.08) | 1.17(1.13,1.22) | 31,490(2.2) | 24.0 | 0.96(0.94,0.99) | 1.24(1.20,1.27) |
| Serious Mental Illness | 20,065(2.5) | 25.7 | 1.33(1.29,1.38) | 1.23(1.19,1.27) | 28,420(2.0) | 31.1 | 1.38(1.35,1.42) | 1.22(1.19,1.25) |
| Asthma | 155,055(19.5) | 22.4 | 1.13(1.11,1.14) | 1.07(1.05,1.08) | 279,720(19.5) | 26.3 | 1.11(1.10,1.12) | 1.03(1.02,1.04) |
| COPD | 66,110(8.3) | 22.2 | 1.10(1.08,1.12) | 1.13(1.10,1.15) | 130,170(9.1) | 24.9 | 1.01(1.00,1.02) | 1.10(1.09,1.12) |
| Stroke and TIA | 64,300(8.1) | 20.7 | 1.00(0.98,1.02) | 1.07(1.05,1.09) | 133,430(9.3) | 23.0 | 0.90(0.89,0.91) | 1.04(1.03,1.06) |
| **Diabetic Medication** | | | | | | | | |
| No medication | 193,125(24.3) | 24.0 | 1 | 1 | | | | |
| Non-insulin | 494,220(62.3) | 18.0 | 0.70(0.69,0.71) | 0.68(0.67,0.69) | | | | |
| Insulin | 106,425(13.4) | 27.4 | 1.20(1.18,1.22) | 1.16(1.14,1.18) | | | | |

Rapid weight gain: > 0.5 kilograms/metre$^2$/year. N (%): Total number (N) and percentage (%) from each population strata contributing to the analysis. %: percentage who gained weight rapidly. OR: unadjusted Odds Ratio generated in logistic regression models. aOR: adjusted Odds Ratio generated in logistic regression models adjusted for age, sex, ethnicity, Index of Multiple Deprivation. COPD: Chronic Obstructive Pulmonary Disease. TIA: Transient Ischaemic Attack.

Figure S3. Average rate of weight change before the onset of the COVID-19 pandemic amongst adults living in England with hypertension or type 2 diabetes

Average rate of weight change (δ*) before the onset of the Coronavirus-19 pandemic amongst adults living in England with hypertension or type 2 diabetes

| | Adults with type 2 diabetes | | Adults with hypertension | |
|---|---|---|---|---|
| **Population Subgroup** | **N (%)** | **Median (IQR)** | **N (%)** | **Median (IQR)** |
| All | 820,390 (100.0) | -0.1 (-0.6, 0.3) | 1,637,165 (100.0) | 0.0 (-0.5, 0.4) |
| Sex | | | | |
| Female | 362,225 (44.2) | -0.1 (-0.6, 0.4) | 815,600 (49.8) | 0.0 (-0.5, 0.5) |
| Male | 458,160 (55.8) | -0.1 (-0.5, 0.3) | 821,565 (50.2) | 0.0 (-0.4, 0.4) |
| Age in years | | | | |
| 18-29 | 2,700 (0.3) | 0.2 (-0.7, 1.2) | 4,725 (0.3) | 0.4 (-0.3, 1.2) |
| 30-39 | 17,955 (2.2) | 0.0 (-0.7, 0.7) | 25,460 (1.6) | 0.2 (-0.4, 0.9) |
| 40-49 | 64,845 (7.9) | -0.1 (-0.6, 0.5) | 96,910 (5.9) | 0.1 (-0.4, 0.7) |
| 50-59 | 161,545 (19.7) | -0.1 (-0.6, 0.4) | 277,865 (17.0) | 0.1 (-0.4, 0.6) |
| 60-69 | 222,530 (27.1) | -0.1 (-0.6, 0.4) | 418,340 (25.6) | 0.0 (-0.4, 0.5) |
| 70-79 | 226,065 (27.6) | -0.1 (-0.5, 0.3) | 508,085 (31.0) | -0.0 (-0.5, 0.3) |
| 80-90 | 124,750 (15.2) | -0.2 (-0.6, 0.2) | 305,775 (18.7) | -0.1 (-0.5, 0.3) |
| Ethnicity | | | | |
| White | 667,300 (81.3) | -0.1 (-0.6, 0.4) | 1,466,955 (89.6) | 0.0 (-0.5, 0.4) |
| Black | 23,675 (2.9) | -0.1 (-0.5, 0.4) | 35,155 (2.1) | 0.0 (-0.5, 0.4) |
| South Asian | 90,590 (11.0) | -0.1 (-0.5, 0.3) | 88,040 (5.4) | 0.0 (-0.4, 0.3) |
| Mixed | 7,590 (0.9) | -0.1 (-0.6, 0.4) | 10,970 (0.7) | 0.0 (-0.5, 0.4) |
| Chinese and other | 31,230 (3.8) | -0.1 (-0.5, 0.3) | 36,045 (2.2) | 0.0 (-0.4, 0.4) |
| Index of Multiple Deprivation | | | | |
| 1 (most deprived) | 199,200 (24.3) | -0.1 (-0.6, 0.4) | 328,465 (20.1) | 0.0 (-0.5, 0.5) |
| 2 | 179,480 (21.9) | -0.1 (-0.6, 0.4) | 329,350 (20.1) | 0.0 (-0.5, 0.4) |
| 3 | 173,775 (21.2) | -0.1 (-0.6, 0.3) | 358,635 (21.9) | 0.0 (-0.5, 0.4) |
| 4 | 147,370 (18.0) | -0.1 (-0.6, 0.3) | 329,895 (20.2) | 0.0 (-0.4, 0.4) |
| 5 (least deprived) | 120,565 (14.7) | -0.1 (-0.6, 0.3) | 290,820 (17.8) | 0.0 (-0.4, 0.4) |
| Comorbid Long Term Condition | | | | |
| Hypertension | 550,775 (67.1) | -0.1 (-0.6, 0.3) | | |
| Type 2 Diabetes | | | 550,775 (33.6) | -0.1 (-0.6, 0.3) |
| Type 1 Diabetes | | | 22,575 (1.4) | 0.1 (-0.3, 0.5) |
| Chronic Cardiac Disease | 195,785 (23.9) | -0.1 (-0.6, 0.3) | 380,610 (23.2) | -0.0 (-0.5, 0.4) |
| Learning Disability | 7,020 (0.9) | -0.1 (-0.7, 0.5) | 10,090 (0.6) | 0.0 (-0.6, 0.6) |
| Depression | 218,780 (26.7) | -0.1 (-0.6, 0.4) | 411,930 (25.2) | 0.0 (-0.5, 0.5) |
| Serious Mental Illness | 20,120 (2.5) | -0.1 (-0.7, 0.5) | 28,720 (1.8) | 0.0 (-0.6, 0.6) |
| Dementia | 20,695 (2.5) | -0.2 (-0.7, 0.3) | 39,425 (2.4) | -0.1 (-0.7, 0.4) |
| Asthma | 163,700 (20.0) | -0.1 (-0.6, 0.4) | 334,025 (20.4) | 0.0 (-0.5, 0.5) |
| Chronic Obstructive Pulmonary Disease | 72,010 (8.8) | -0.1 (-0.6, 0.4) | 160,230 (9.8) | 0.0 (-0.5, 0.5) |
| Stroke and TIA | 70,615 (8.6) | -0.1 (-0.6, 0.3) | 156,060 (9.5) | -0.1 (-0.5, 0.4) |
| Diabetes Treatment | | | | |
| No pharmacological treatment | 202,010 (24.6) | -0.1 (-0.6, 0.4) | | |
| Non-insulin | 503,480 (61.4) | -0.1 (-0.6, 0.3) | | |
| Insulin | 114,895 (14.0) | 0.0 (-0.5, 0.5) | | |

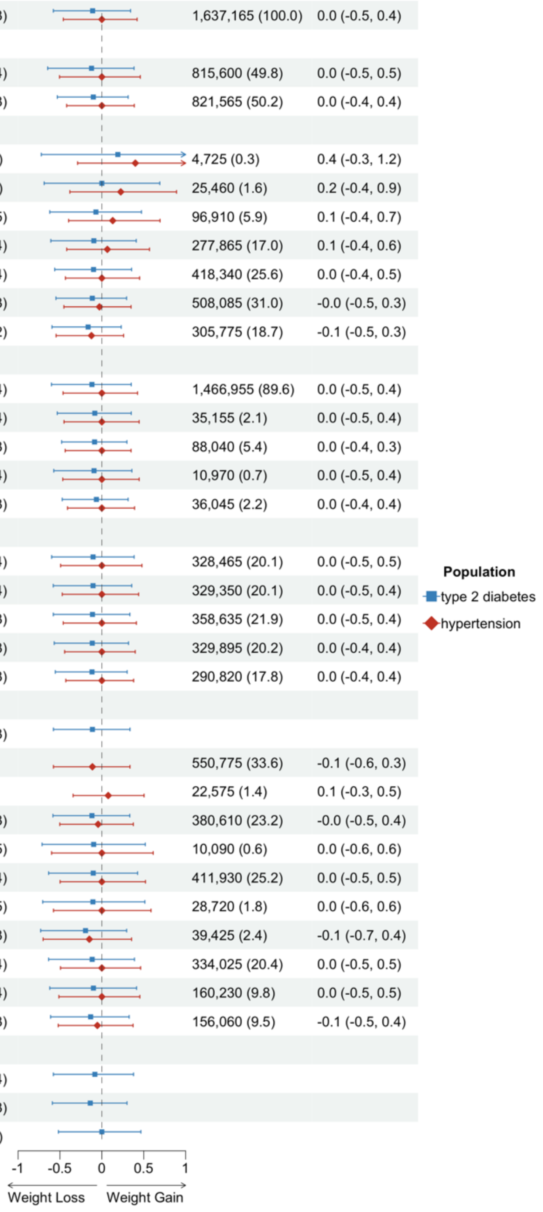

* δ = individual level rate of weight change (in kilograms/metre$^2$/year) calculated from data recorded in general practice healthcare records

Figure S4. Odds of rapid weight gain amongst adults living in England with hypertension before the onset of the COVID-19 pandemic

The rate of weight gain before the onset of the pandemic (δ-prepandemic) was calculated using a random BMI measure from each of from period-0 (March 2015 - February 2018) and period-1 (March 2018 - February 2020) (Supplementary Figure 1).

| | Adults with type 2 diabetes | | | Adults with hypertension | | |
|---|---|---|---|---|---|---|
| **Population Subgroup** | **N (%)** | **Prevalence (%)** | **aOR (95% CI)** | **N (%)** | **Prevalence (%)** | **aOR (95% CI)** |
| All | 820,390 (100.0) | 19.2 | | 1,637,165 (100.0) | 21.8 | |
| Sex | | | | | | |
| Female | 362,225 (44.2) | 21.1 | 1 | 815,600 (49.8) | 23.5 | 1 |
| Male | 458,160 (55.8) | 17.8 | 0.81 (0.80 to 0.82) | 821,565 (50.2) | 20.2 | 0.80 (0.80 to 0.81) |
| Age in years | | | | | | |
| 18-29 | 2,700 (0.3) | 40.9 | 1 | 4,725 (0.3) | 46.6 | 1 |
| 30-39 | 17,955 (2.2) | 30.6 | 0.67 (0.62 to 0.73) | 25,460 (1.6) | 38.8 | 0.75 (0.71 to 0.80) |
| 40-49 | 64,845 (7.9) | 24.1 | 0.50 (0.46 to 0.54) | 96,910 (5.9) | 32.3 | 0.59 (0.55 to 0.62) |
| 50-59 | 161,545 (19.7) | 21.9 | 0.43 (0.40 to 0.47) | 277,865 (17.0) | 27.7 | 0.47 (0.44 to 0.49) |
| 60-69 | 222,530 (27.1) | 19.7 | 0.37 (0.35 to 0.41) | 418,340 (25.6) | 23.0 | 0.36 (0.34 to 0.38) |
| 70-79 | 226,065 (27.6) | 17.0 | 0.31 (0.28 to 0.33) | 508,085 (31.0) | 18.5 | 0.27 (0.26 to 0.29) |
| 80-90 | 124,750 (15.2) | 14.5 | 0.25 (0.23 to 0.27) | 305,775 (18.7) | 15.4 | 0.21 (0.20 to 0.23) |
| Ethnicity | | | | | | |
| White | 667,300 (81.3) | 19.7 | 1 | 1,466,955 (89.6) | 22.0 | 1 |
| Black | 23,675 (2.9) | 19.2 | 0.83 (0.80 to 0.86) | 35,155 (2.1) | 22.8 | 0.81 (0.79 to 0.84) |
| South Asian | 90,590 (11.0) | 16.8 | 0.70 (0.68 to 0.71) | 88,040 (5.4) | 18.7 | 0.67 (0.66 to 0.68) |
| Chinese and other | 31,230 (3.8) | 17.3 | 0.74 (0.72 to 0.77) | 36,045 (2.2) | 20.3 | 0.75 (0.73 to 0.77) |
| Mixed | 7,590 (0.9) | 20.0 | 0.87 (0.82 to 0.92) | 10,970 (0.7) | 23.2 | 0.84 (0.81 to 0.88) |
| Index of Multiple Deprivation | | | | | | |
| 1 (most deprived) | 199,200 (24.3) | 21.0 | 1 | 328,465 (20.1) | 24.2 | 1 |
| 2 | 179,480 (21.9) | 19.8 | 0.96 (0.94 to 0.97) | 329,350 (20.1) | 22.7 | 0.95 (0.94 to 0.96) |
| 3 | 173,775 (21.2) | 18.8 | 0.91 (0.90 to 0.93) | 358,635 (21.9) | 21.4 | 0.91 (0.90 to 0.92) |
| 4 | 147,370 (18.0) | 18.3 | 0.89 (0.87 to 0.90) | 329,895 (20.2) | 20.9 | 0.89 (0.88 to 0.90) |
| 5 (least deprived) | 120,565 (14.7) | 17.4 | 0.84 (0.83 to 0.86) | 290,820 (17.8) | 19.9 | 0.84 (0.83 to 0.86) |
| Long Term Condition | | | | | | |
| Hypertension | 550,775 (67.1) | 18.9 | 1.08 (1.07 to 1.09) | | | |
| Diabetes Type 2 | | | | 550,775 (33.6) | 18.9 | 0.81 (0.81 to 0.82) |
| Diabetes Type 1 | | | | 22,575 (1.4) | 25.2 | 0.98 (0.95 to 1.01) |
| Chronic Cardiac Disease | 195,785 (23.9) | 18.8 | 1.13 (1.11 to 1.14) | 380,610 (23.2) | 20.1 | 1.07 (1.06 to 1.08) |
| Learning Disability | 7,020 (0.9) | 25.5 | 1.11 (1.05 to 1.17) | 10,090 (0.6) | 29.1 | 1.09 (1.04 to 1.14) |
| Depression | 218,780 (26.7) | 22.6 | 1.17 (1.15 to 1.18) | 411,930 (25.2) | 25.8 | 1.17 (1.16 to 1.18) |
| Dementia | 20,695 (2.5) | 18.0 | 1.14 (1.10 to 1.18) | 39,425 (2.4) | 19.9 | 1.19 (1.16 to 1.22) |
| Serious Mental Illness | 20,120 (2.5) | 25.3 | 1.26 (1.22 to 1.31) | 28,720 (1.8) | 27.8 | 1.20 (1.17 to 1.23) |
| Asthma | 163,700 (20.0) | 21.2 | 1.09 (1.07 to 1.10) | 334,025 (20.4) | 23.6 | 1.06 (1.05 to 1.07) |
| COPD | 72,010 (8.8) | 22.0 | 1.29 (1.26 to 1.31) | 160,230 (9.8) | 23.2 | 1.23 (1.22 to 1.25) |
| Stroke and TIA | 70,615 (8.6) | 18.7 | 1.09 (1.07 to 1.11) | 156,060 (9.5) | 20.0 | 1.05 (1.04 to 1.07) |
| Diabetic Medication | | | | | | |
| Non pharmacological treatment | 202,010 (24.6) | 20.4 | 1 | | | |
| Non-Insulin | 503,480 (61.4) | 17.8 | 0.80 (0.79 to 0.81) | | | |
| Insulin | 114,895 (14.0) | 23.7 | 1.14 (1.12 to 1.16) | | | |

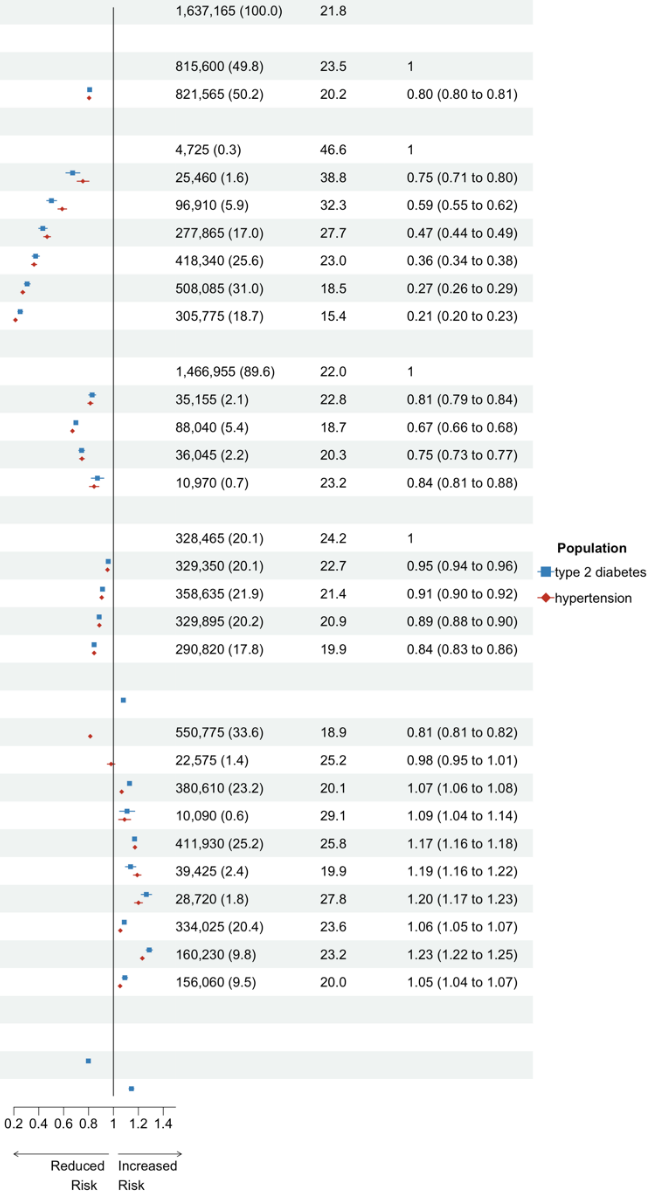

Rapid weight gain: > 0.5 kilograms/metre$^2$/year. N (%): Total number (N) and percentage (%) from each population strata contributing to the analysis. Prevalence (%): percentage who gained weight rapidly. aOR: adjusted Odds Ratio generated in logistic regression models adjusted for age, sex, ethnicity, and Index of Multiple Deprivation. 95% CI: 95% Confidence Interval of aOR. COPD: Chronic Obstructive Pulmonary Disease. TIA: Transient Ischaemic Attack

Figure S5. Sex stratified odds of rapid weight gain amongst adults living in England with type 2 diabetes during the COVID-19 pandemic.

| Population Subgroup | Female N (%) | Female Prevalence (%) | Female aOR (95% CI) | Male N (%) | Male Prevalence (%) | Male aOR (95% CI) |
|---|---|---|---|---|---|---|
| All | 345,050 (100.0) | 23.0 | | 448,725 (100.0) | 19.0 | |
| **Age in Years** | | | | | | |
| 18-29 | 2,265 (0.7) | 30.7 | 1 | 1,050 (0.2) | 29.0 | 1 |
| 30-39 | 11,210 (3.2) | 28.2 | 0.91 (0.82 to 1.00) | 8,580 (1.9) | 22.0 | 0.69 (0.60 to 0.80) |
| 40-49 | 31,020 (9.0) | 25.0 | 0.77 (0.71 to 0.85) | 37,390 (8.3) | 20.6 | 0.64 (0.56 to 0.73) |
| 50-59 | 67,470 (19.6) | 24.8 | 0.74 (0.68 to 0.81) | 95,580 (21.3) | 20.4 | 0.61 (0.53 to 0.70) |
| 60-69 | 88,080 (25.5) | 23.6 | 0.69 (0.63 to 0.76) | 130,250 (29.0) | 19.7 | 0.58 (0.51 to 0.66) |
| 70-79 | 91,580 (26.5) | 21.8 | 0.61 (0.56 to 0.67) | 123,325 (27.5) | 17.8 | 0.51 (0.44 to 0.58) |
| 80-90 | 53,420 (15.5) | 18.9 | 0.51 (0.47 to 0.56) | 52,545 (11.7) | 15.9 | 0.44 (0.39 to 0.51) |
| **Ethnicity** | | | | | | |
| White | 272,505 (79.0) | 23.7 | 1 | 367,685 (81.9) | 19.5 | 1 |
| Black | 12,245 (3.5) | 24.4 | 0.96 (0.92 to 1.00) | 11,685 (2.6) | 19.9 | 0.94 (0.89 to 0.98) |
| South Asian | 42,030 (12.2) | 18.9 | 0.68 (0.67 to 0.70) | 47,720 (10.6) | 16.0 | 0.72 (0.70 to 0.74) |
| Chinese and Other | 14,450 (4.2) | 19.2 | 0.71 (0.68 to 0.74) | 17,730 (4.0) | 16.7 | 0.77 (0.74 to 0.80) |
| Mixed | 3,820 (1.1) | 25.0 | 0.99 (0.92 to 1.07) | 3,895 (0.9) | 20.3 | 0.97 (0.90 to 1.05) |
| **Index of Multiple Deprivation** | | | | | | |
| IMD 1 (most deprived) | 90,020 (26.1) | 23.9 | 1 | 101,750 (22.7) | 20.3 | 1 |
| IMD 2 | 77,565 (22.5) | 23.1 | 0.97 (0.95 to 0.99) | 96,545 (21.5) | 19.4 | 0.96 (0.94 to 0.98) |
| IMD 3 | 71,830 (20.8) | 22.9 | 0.96 (0.93 to 0.98) | 97,035 (21.6) | 18.9 | 0.92 (0.90 to 0.94) |
| IMD 4 | 58,685 (17.0) | 22.3 | 0.92 (0.90 to 0.94) | 83,900 (18.7) | 18.4 | 0.89 (0.87 to 0.91) |
| IMD 5 (least deprived) | 46,950 (13.6) | 21.7 | 0.89 (0.87 to 0.92) | 69,490 (15.5) | 17.6 | 0.85 (0.83 to 0.87) |
| **Comorbid Long Term Condition** | | | | | | |
| Hypertension | 225,885 (65.5) | 22.6 | 1.04 (1.02 to 1.06) | 294,480 (65.6) | 19.2 | 1.09 (1.07 to 1.11) |
| Chronic Cardiac Disease | 61,310 (17.8) | 22.9 | 1.08 (1.06 to 1.11) | 120,655 (26.9) | 19.4 | 1.10 (1.08 to 1.12) |
| Learning Disability | 3,080 (0.9) | 26.5 | 1.04 (0.95 to 1.12) | 3,950 (0.9) | 24.6 | 1.21 (1.12 to 1.30) |
| Depression | 115,450 (33.5) | 25.2 | 1.12 (1.11 to 1.14) | 93,325 (20.8) | 21.4 | 1.14 (1.12 to 1.16) |
| Serious Mental Illness | 9,795 (2.8) | 27.4 | 1.21 (1.15 to 1.26) | 10,270 (2.3) | 24.1 | 1.26 (1.21 to 1.32) |
| Dementia | 8,645 (2.5) | 22.8 | 1.16 (1.10 to 1.22) | 8,695 (1.9) | 20.0 | 1.20 (1.13 to 1.26) |
| Asthma | 82,805 (24.0) | 24.3 | 1.06 (1.04 to 1.08) | 72,250 (16.1) | 20.2 | 1.07 (1.05 to 1.09) |
| COPD | 27,650 (8.0) | 24.4 | 1.09 (1.06 to 1.13) | 38,460 (8.6) | 20.7 | 1.15 (1.12 to 1.18) |
| Stroke and TIA | 26,040 (7.5) | 22.7 | 1.06 (1.03 to 1.09) | 38,265 (8.5) | 19.3 | 1.08 (1.05 to 1.11) |
| **Diabetes Treatment** | | | | | | |
| Non Pharmacological Treatment | 91,290 (26.5) | 26.4 | 1 | 101,835 (22.7) | 21.8 | 1 |
| Non Insulin | 205,950 (59.7) | 19.9 | 0.66 (0.65 to 0.67) | 288,275 (64.2) | 16.7 | 0.70 (0.69 to 0.71) |
| Insulin | 47,810 (13.9) | 29.6 | 1.11 (1.08 to 1.14) | 58,615 (13.1) | 25.6 | 1.20 (1.17 to 1.23) |

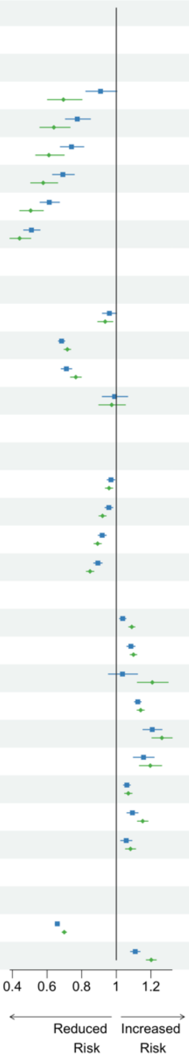

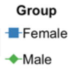

Rapid weight gain: > 0.5 kilograms/metre$^2$/year. N (%): Total number (N) and percentage (%) from each population strata contributing to the analysis. Prevalence (%): percentage who gained weight rapidly. aOR: adjusted Odds Ratio generated in logistic regression models adjusted for age, ethnicity, and Index of Multiple Deprivation. 95% CI: 95% Confidence Interval of aOR. COPD: Chronic Obstructive Pulmonary Disease. TIA: Transient Ischaemic Attack.

Figure S6. Ethnicity stratified odds of rapid weight gain amongst adults living in England with type 2 diabetes during the COVID-19 pandemic.

| | South Asian | | | Black | | |
|---|---|---|---|---|---|---|
| Population Subgroup | N (%) | Prevalence (%) | aOR (95% CI) | N (%) | Prevalence (%) | aOR (95% CI) |
| All | 89,750 (100.0) | 17.3 | | 23,935 (100.0) | 22.2 | |
| Sex | | | | | | |
| Female | 42,030 (46.8) | 18.9 | 1 | 12,245 (51.2) | 24.4 | 1 |
| Male | 47,720 (53.2) | 16.0 | 0.83 (0.80 to 0.86) | 11,685 (48.8) | 19.9 | 0.77 (0.72 to 0.82) |
| Age in Years | | | | | | |
| 18-29 | 610 (0.7) | 25.4 | 1 | 140 (0.6) | 42.9 | 1 |
| 30-39 | 4,895 (5.5) | 21.8 | 0.83 (0.68 to 1.01) | 925 (3.9) | 29.2 | 0.61 (0.42 to 0.88) |
| 40-49 | 16,580 (18.5) | 18.4 | 0.69 (0.57 to 0.83) | 3,625 (15.1) | 23.4 | 0.47 (0.33 to 0.66) |
| 50-59 | 22,425 (25.0) | 17.1 | 0.63 (0.53 to 0.77) | 7,630 (31.9) | 22.6 | 0.45 (0.32 to 0.63) |
| 60-69 | 25,525 (28.4) | 16.1 | 0.59 (0.49 to 0.71) | 6,110 (25.5) | 21.4 | 0.42 (0.30 to 0.59) |
| 70-79 | 13,685 (15.2) | 16.9 | 0.62 (0.52 to 0.75) | 3,130 (13.1) | 20.4 | 0.38 (0.27 to 0.54) |
| 80-90 | 6,025 (6.7) | 17.1 | 0.63 (0.52 to 0.76) | 2,375 (9.9) | 19.4 | 0.36 (0.25 to 0.51) |
| Index of Multiple Deprivation | | | | | | |
| IMD 1 (most deprived) | 34,165 (38.1) | 18.2 | 1 | 10,240 (42.8) | 23.1 | 1 |
| IMD 2 | 25,875 (28.8) | 17.2 | 0.95 (0.91 to 0.99) | 6,375 (26.6) | 21.9 | 0.94 (0.87 to 1.01) |
| IMD 3 | 15,840 (17.6) | 17.1 | 0.95 (0.90 to 1.00) | 3,865 (16.1) | 22.0 | 0.96 (0.88 to 1.05) |
| IMD 4 | 8,210 (9.1) | 15.9 | 0.87 (0.82 to 0.93) | 2,225 (9.3) | 20.0 | 0.85 (0.76 to 0.95) |
| IMD 5 (least deprived) | 5,655 (6.3) | 15.6 | 0.86 (0.80 to 0.93) | 1,220 (5.1) | 21.3 | 0.93 (0.80 to 1.07) |
| Comorbid Long Term Condition | | | | | | |
| Hypertension | 51,035 (56.9) | 17.5 | 1.10 (1.06 to 1.14) | 15,275 (63.8) | 22.2 | 1.10 (1.02 to 1.17) |
| Chronic Cardiac Disease | 17,725 (19.7) | 18.2 | 1.18 (1.13 to 1.23) | 2,990 (12.5) | 23.1 | 1.16 (1.05 to 1.27) |
| Learning Disability | 485 (0.5) | 28.9 | 1.69 (1.38 to 2.06) | 130 (0.5) | 23.1 | 0.95 (0.62 to 1.41) |
| Depression | 15,820 (17.6) | 19.4 | 1.15 (1.10 to 1.20) | 4,105 (17.2) | 23.9 | 1.07 (0.98 to 1.16) |
| Serious Mental Illness | 2,630 (2.9) | 22.2 | 1.33 (1.21 to 1.46) | 1,125 (4.7) | 25.3 | 1.15 (1.00 to 1.32) |
| Dementia | 1,775 (2.0) | 19.2 | 1.16 (1.03 to 1.31) | 560 (2.3) | 23.2 | 1.18 (0.95 to 1.44) |
| Asthma | 16,210 (18.1) | 18.3 | 1.07 (1.02 to 1.12) | 3,760 (15.7) | 24.3 | 1.13 (1.04 to 1.23) |
| COPD | 2,935 (3.3) | 18.2 | 1.14 (1.04 to 1.26) | 650 (2.7) | 23.8 | 1.20 (0.99 to 1.44) |
| Stroke and TIA | 5,130 (5.7) | 19.3 | 1.21 (1.13 to 1.30) | 1,365 (5.7) | 22.3 | 1.10 (0.96 to 1.26) |
| Diabetes Treatment | | | | | | |
| Non Pharmacological | 15,800 (17.6) | 60.5 | 1 | 5,210 (21.8) | 25.0 | 1 |
| Non Insulin | 63,220 (70.4) | 5.2 | 0.68 (0.65 to 0.71) | 15,405 (64.4) | 20.1 | 0.76 (0.70 to 0.82) |
| Insulin | 10,735 (12.0) | 25.1 | 1.27 (1.20 to 1.35) | 3,315 (13.9) | 27.6 | 1.16 (1.05 to 1.28) |

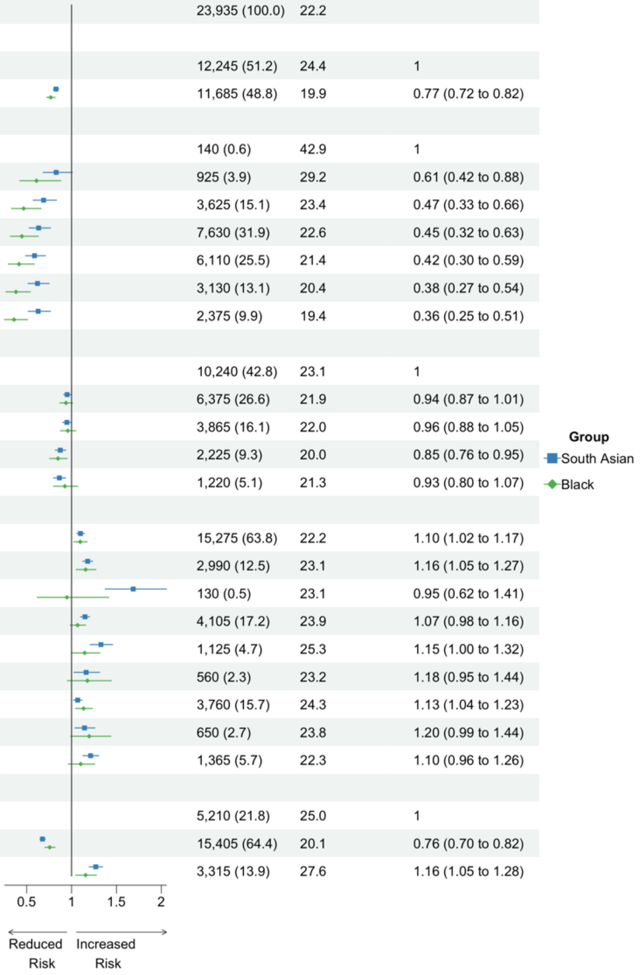

Rapid weight gain: > 0.5 kilograms/metre$^2$/year. N (%): Total number (N) and percentage (%) from each population strata contributing to the analysis. Prevalence (%): percentage who gained weight rapidly. aOR: adjusted Odds Ratio generated in logistic regression models adjusted for age, sex, and Index of Multiple Deprivation. 95% CI: 95% Confidence Interval of aOR. COPD: Chronic Obstructive Pulmonary Disease. TIA: Transient Ischaemic Attack

Figure S7. Age stratified odds of rapid weight gain amongst adults living in England with type 2 diabetes during the COVID-19 pandemic.

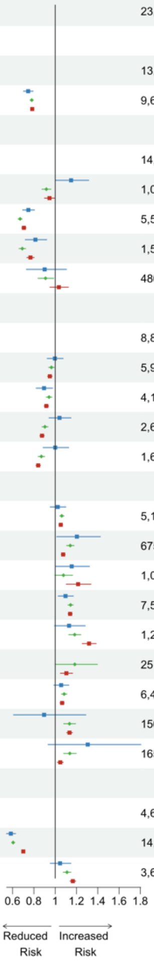

| Population Subgroup | 18-39 years N (%) | 18-39 years Prevalence (%) | 18-39 years aOR (95% CI) | 40-59 years N (%) | 40-59 years Prevalence (%) | 40-59 years aOR (95% CI) | 60-79 years N (%) | 60-79 years Prevalence (%) | 60-79 years aOR (95% CI) |
|---|---|---|---|---|---|---|---|---|---|
| All | 23,100 (100.0) | 26.2 | | 231,465 (100.0) | 22.3 | | 231,465 (100.0) | 11.9 | |
| Sex | | | | | | | | | |
| Female | 13,475 (58.3) | 28.6 | 1 | 98,490 (42.6) | 24.9 | 1 | 98,490 (41.5) | 13.6 | 1 |
| Male | 9,625 (41.7) | 22.8 | 0.75 (0.70 to 0.79) | 132,975 (57.4) | 20.5 | 0.78 (0.76 to 0.80) | 132,975 (58.5) | 10.7 | 0.78 (0.76 to 0.80) |
| Ethnicity | | | | | | | | | |
| White | 14,485 (62.7) | 27.8 | 1 | 163,365 (70.6) | 23.8 | 1 | 163,365 (84.8) | 10.6 | 1 |
| Black | 1,070 (4.6) | 30.4 | 1.15 (1.00 to 1.31) | 11,255 (4.9) | 22.9 | 0.92 (0.88 to 0.96) | 11,255 (2.1) | 27.9 | 0.92 (0.88 to 0.96) |
| South Asian | 5,505 (23.8) | 22.2 | 0.75 (0.69 to 0.81) | 39,005 (16.9) | 17.7 | 0.67 (0.65 to 0.69) | 39,005 (9.1) | 17.6 | 0.67 (0.65 to 0.69) |
| Chinese and Other | 1,560 (6.8) | 23.4 | 0.81 (0.72 to 0.92) | 14,385 (6.2) | 17.9 | 0.69 (0.66 to 0.72) | 14,385 (3.3) | 18.1 | 0.69 (0.66 to 0.72) |
| Mixed | 480 (2.1) | 26.0 | 0.90 (0.73 to 1.11) | 3,455 (1.5) | 22.6 | 0.91 (0.84 to 0.99) | 3,455 (0.7) | 24.1 | 0.91 (0.84 to 0.99) |
| Index of Multiple Deprivation | | | | | | | | | |
| IMD 1 (most deprived) | 8,850 (38.3) | 26.4 | 1 | 70,710 (30.5) | 23.0 | 1 | 70,710 (21.8) | 17.2 | 1 |
| IMD 2 | 5,910 (25.6) | 26.2 | 1.00 (0.92 to 1.07) | 55,275 (23.9) | 22.3 | 0.96 (0.94 to 0.99) | 55,275 (21.4) | 13.3 | 0.96 (0.94 to 0.99) |
| IMD 3 | 4,110 (17.8) | 24.6 | 0.90 (0.82 to 0.98) | 45,645 (19.7) | 22.3 | 0.94 (0.92 to 0.97) | 45,645 (21.9) | 10.7 | 0.94 (0.92 to 0.97) |
| IMD 4 | 2,600 (11.3) | 27.7 | 1.04 (0.94 to 1.15) | 34,310 (14.8) | 21.9 | 0.90 (0.88 to 0.93) | 34,310 (19.1) | 9.1 | 0.90 (0.88 to 0.93) |
| IMD 5 (least deprived) | 1,635 (7.1) | 26.9 | 1.00 (0.89 to 1.13) | 25,520 (11.0) | 21.4 | 0.87 (0.84 to 0.90) | 25,520 (15.9) | 7.9 | 0.87 (0.84 to 0.90) |
| Comorbid Long Term Condition | | | | | | | | | |
| Hypertension | 5,100 (22.1) | 26.3 | 1.02 (0.95 to 1.10) | 116,155 (50.2) | 23.0 | 1.06 (1.04 to 1.08) | 116,155 (71.9) | 8.6 | 1.06 (1.04 to 1.08) |
| Chronic Cardiac Disease | 675 (2.9) | 29.6 | 1.20 (1.01 to 1.42) | 26,455 (11.4) | 24.1 | 1.14 (1.11 to 1.18) | 26,455 (26.1) | 5.6 | 1.14 (1.11 to 1.18) |
| Learning Disability | 1,075 (4.7) | 28.8 | 1.15 (1.01 to 1.32) | 3,525 (1.5) | 24.8 | 1.08 (1.00 to 1.16) | 3,525 (0.5) | 37.9 | 1.08 (1.00 to 1.16) |
| Depression | 7,570 (32.8) | 28.7 | 1.10 (1.03 to 1.17) | 74,815 (32.3) | 25.1 | 1.14 (1.12 to 1.17) | 74,815 (25.1) | 17.3 | 1.14 (1.12 to 1.17) |
| Serious Mental Illness | 1,270 (5.5) | 28.0 | 1.13 (0.99 to 1.28) | 9,305 (4.0) | 25.8 | 1.18 (1.13 to 1.24) | 9,305 (2.0) | 28.4 | 1.18 (1.13 to 1.24) |
| Dementia | 25 (0.1) | | Redacted | 730 (0.3) | 25.3 | 1.18 (1.00 to 1.40) | 730 (2.0) | 2.1 | 1.18 (1.00 to 1.40) |
| Asthma | 6,415 (27.8) | 27.7 | 1.06 (0.99 to 1.13) | 49,055 (21.2) | 24.4 | 1.08 (1.06 to 1.11) | 49,055 (18.7) | 14.7 | 1.08 (1.06 to 1.11) |
| COPD | 150 (0.6) | 23.3 | 0.90 (0.61 to 1.29) | 8,835 (3.8) | 25.9 | 1.13 (1.08 to 1.19) | 8,835 (10.3) | 5.2 | 1.13 (1.08 to 1.19) |
| Stroke and TIA | 165 (0.7) | 33.3 | 1.31 (0.93 to 1.80) | 8,020 (3.5) | 25.0 | 1.14 (1.08 to 1.20) | 8,020 (8.9) | 5.2 | 1.14 (1.08 to 1.20) |
| Diabetes Treatment | | | | | | | | | |
| Non Pharmacological Treatment | 4,685 (20.3) | 32.7 | 1 | 44,265 (19.1) | 28.2 | 1 | 44,265 (24.4) | 11.8 | 1 |
| Non Insulin | 14,800 (64.1) | 22.1 | 0.58 (0.54 to 0.63) | 157,645 (68.1) | 19.1 | 0.60 (0.59 to 0.62) | 157,645 (61.7) | 11.2 | 0.60 (0.59 to 0.62) |
| Insulin | 3,610 (15.6) | 34.8 | 1.05 (0.95 to 1.15) | 29,555 (12.8) | 31.0 | 1.11 (1.08 to 1.15) | 29,555 (13.9) | 15.2 | 1.11 (1.08 to 1.15) |

Rapid weight gain: > 0.5 kilograms/metre$^2$/year. N (%): Total number (N) and percentage (%) from each population strata contributing to the analysis. Prevalence (%): percentage who gained weight rapidly. aOR: adjusted Odds Ratio generated in logistic regression models adjusted for sex, ethnicity, and Index of Multiple Deprivation. 95% CI: 95% Confidence Interval of aOR. COPD: Chronic Obstructive Pulmonary Disease. TIA: Transient Ischaemic Attack.

Figure S8. Sex stratified odds of rapid weight gain amongst adults living in England with hypertension during the COVID-19 pandemic.

| Population Subgroup | Female N (%) | Female Prevalence (%) | Female aOR (95% CI) | Male N (%) | Male Prevalence (%) | Male aOR (95% CI) |
|---|---|---|---|---|---|---|
| All | 700,230 (100.0) | 27.2 | | 735,840 (100.0) | 22.4 | |
| Age in Years | | | | | | |
| 18-29 | 3,175 (0.5) | 45.7 | 1 | 1,550 (0.2) | 40.0 | 1 |
| 30-39 | 13,605 (1.9) | 41.9 | 0.88 (0.81 to 0.95) | 9,625 (1.3) | 35.0 | 0.82 (0.73 to 0.91) |
| 40-49 | 44,235 (6.3) | 37.1 | 0.73 (0.68 to 0.78) | 45,165 (6.1) | 29.7 | 0.65 (0.58 to 0.72) |
| 50-59 | 114,795 (16.4) | 33.4 | 0.61 (0.57 to 0.65) | 138,315 (18.8) | 27.3 | 0.56 (0.51 to 0.62) |
| 60-69 | 168,340 (24.0) | 28.8 | 0.49 (0.46 to 0.53) | 210,175 (28.6) | 23.4 | 0.46 (0.41 to 0.50) |
| 70-79 | 218,460 (31.2) | 24.1 | 0.38 (0.35 to 0.41) | 227,660 (30.9) | 19.1 | 0.35 (0.32 to 0.39) |
| 80-90 | 137,615 (19.7) | 19.8 | 0.29 (0.27 to 0.32) | 103,350 (14.0) | 16.4 | 0.29 (0.26 to 0.32) |
| Ethnicity | | | | | | |
| White | 619,810 (88.5) | 27.5 | 1 | 657,435 (89.3) | 22.7 | 1 |
| Black | 18,195 (2.6) | 29.7 | 0.90 (0.87 to 0.93) | 14,220 (1.9) | 24.0 | 0.87 (0.84 to 0.91) |
| South Asian | 39,890 (5.7) | 22.0 | 0.62 (0.61 to 0.64) | 41,785 (5.7) | 18.9 | 0.67 (0.65 to 0.69) |
| Chinese and Other | 16,930 (2.4) | 23.2 | 0.68 (0.65 to 0.70) | 17,765 (2.4) | 20.0 | 0.72 (0.70 to 0.75) |
| Mixed | 5,405 (0.8) | 30.3 | 0.94 (0.88 to 0.99) | 4,635 (0.6) | 23.8 | 0.89 (0.83 to 0.95) |
| Index of Multiple Deprivation | | | | | | |
| IMD 1 (most deprived) | 147,095 (21.0) | 29.6 | 1 | 143,735 (19.5) | 25.1 | 1 |
| IMD 2 | 143,480 (20.5) | 28.0 | 0.95 (0.94 to 0.97) | 146,805 (20.0) | 23.4 | 0.94 (0.92 to 0.96) |
| IMD 3 | 152,905 (21.8) | 26.8 | 0.91 (0.90 to 0.93) | 162,625 (22.1) | 22.2 | 0.89 (0.88 to 0.91) |
| IMD 4 | 137,940 (19.7) | 26.1 | 0.88 (0.87 to 0.90) | 149,785 (20.4) | 21.2 | 0.84 (0.83 to 0.86) |
| IMD 5 (least deprived) | 118,810 (17.0) | 25.0 | 0.85 (0.83 to 0.86) | 132,890 (18.1) | 20.0 | 0.79 (0.77 to 0.80) |
| Comorbid Long Term Condition | | | | | | |
| Type 2 Diabetes | 225,885 (32.3) | 22.6 | 0.73 (0.73 to 0.74) | 294,480 (40.0) | 19.2 | 0.75 (0.74 to 0.76) |
| Type 1 Diabetes | 8,120 (1.2) | 32.1 | 1.07 (1.02 to 1.13) | 12,000 (1.6) | 26.6 | 1.05 (1.01 to 1.10) |
| Chronic Cardiac Disease | 127,230 (18.2) | 25.3 | 1.05 (1.03 to 1.06) | 202,500 (27.5) | 21.5 | 1.06 (1.05 to 1.08) |
| Learning Disability | 4,145 (0.6) | 32.7 | 0.98 (0.92 to 1.05) | 5,715 (0.8) | 31.3 | 1.21 (1.14 to 1.28) |
| Depression | 215,400 (30.8) | 30.4 | 1.13 (1.12 to 1.15) | 144,025 (19.6) | 26.0 | 1.17 (1.15 to 1.18) |
| Serious Mental Illness | 14,595 (2.1) | 32.3 | 1.18 (1.13 to 1.22) | 13,825 (1.9) | 29.8 | 1.27 (1.22 to 1.32) |
| Dementia | 17,710 (2.5) | 25.3 | 1.22 (1.18 to 1.26) | 13,780 (1.9) | 22.4 | 1.26 (1.21 to 1.32) |
| Asthma | 157,080 (22.4) | 28.5 | 1.03 (1.02 to 1.05) | 122,640 (16.7) | 23.4 | 1.03 (1.02 to 1.05) |
| COPD | 60,395 (8.6) | 27.1 | 1.06 (1.04 to 1.09) | 69,775 (9.5) | 23.0 | 1.14 (1.12 to 1.16) |
| Stroke and TIA | 59,980 (8.6) | 24.9 | 1.03 (1.01 to 1.05) | 73,450 (10.0) | 21.3 | 1.06 (1.04 to 1.08) |

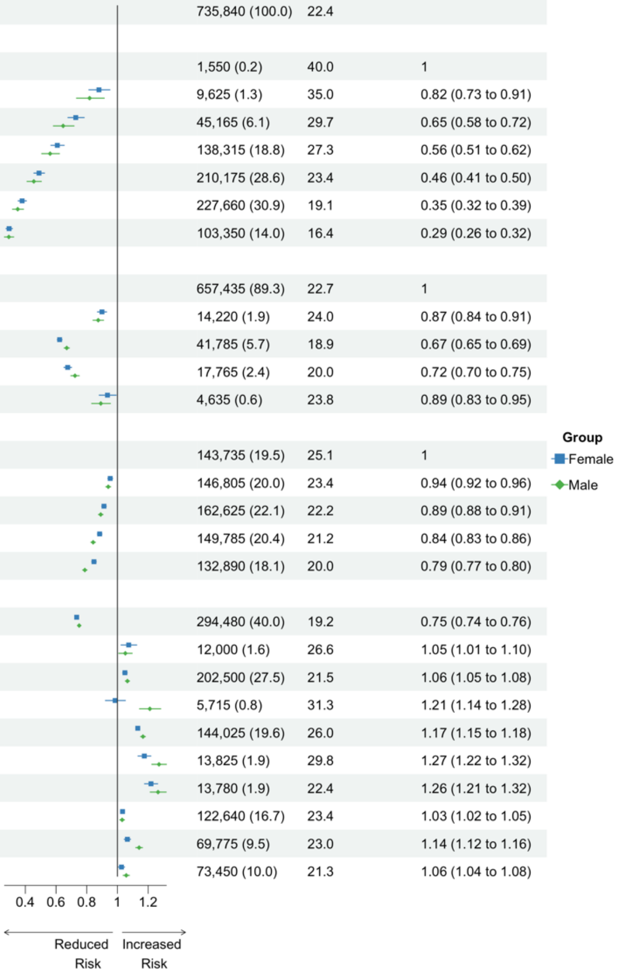

Rapid weight gain: > 0.5 kilograms/metre$^2$/year. N (%): Total number (N) and percentage (%) from each population strata contributing to the analysis. Prevalence (%): percentage who gained weight rapidly. aOR: adjusted Odds Ratio generated in logistic regression models adjusted for age, ethnicity, and Index of Multiple Deprivation. 95% CI: 95% Confidence Interval of aOR. COPD: Chronic Obstructive Pulmonary Disease. TIA: Transient Ischaemic Attack.

Figure S9. Ethnicity stratified odds of rapid weight gain amongst adults living in England with hypertension during the COVID-19 pandemic.

| Population Subgroup | South Asian N (%) | South Asian Prevalence (%) | South Asian aOR (95% CI) | Black N (%) | Black Prevalence (%) | Black aOR (95% CI) |
|---|---|---|---|---|---|---|
| All | 81,670 (100.0) | 20.4 | | 32,415 (100.0) | 27.2 | |
| Sex | | | | | | |
| Female | 39,890 (48.8) | 22.0 | 1 | 18,195 (56.1) | 29.7 | 1 |
| Male | 41,785 (51.2) | 18.9 | 0.82 (0.79 to 0.85) | 14,220 (43.9) | 24.0 | 0.74 (0.70 to 0.78) |
| Age in Years | | | | | | |
| 18-29 | 315 (0.4) | 39.7 | 1 | 125 (0.4) | 44.0 | 1 |
| 30-39 | 2,730 (3.3) | 30.4 | 0.65 (0.51 to 0.83) | 1,050 (3.2) | 39.5 | 0.80 (0.55 to 1.17) |
| 40-49 | 11,170 (13.7) | 24.9 | 0.50 (0.40 to 0.63) | 4,780 (14.7) | 34.7 | 0.66 (0.46 to 0.95) |
| 50-59 | 18,660 (22.8) | 21.7 | 0.42 (0.33 to 0.52) | 10,415 (32.1) | 28.9 | 0.51 (0.36 to 0.74) |
| 60-69 | 25,180 (30.8) | 18.5 | 0.34 (0.27 to 0.43) | 8,175 (25.2) | 24.9 | 0.42 (0.29 to 0.60) |
| 70-79 | 15,835 (19.4) | 18.1 | 0.33 (0.26 to 0.42) | 4,450 (13.7) | 21.9 | 0.34 (0.24 to 0.49) |
| 80-90 | 7,780 (9.5) | 17.4 | 0.31 (0.25 to 0.40) | 3,420 (10.6) | 19.3 | 0.30 (0.21 to 0.43) |
| Index of Multiple Deprivation | | | | | | |
| IMD 1 (most deprived) | 27,940 (34.2) | 21.6 | 1 | 13,565 (41.8) | 28.1 | 1 |
| IMD 2 | 24,290 (29.7) | 20.2 | 0.94 (0.90 to 0.98) | 8,495 (26.2) | 26.6 | 0.93 (0.88 to 0.99) |
| IMD 3 | 15,855 (19.4) | 19.8 | 0.93 (0.89 to 0.98) | 5,460 (16.8) | 27.6 | 1.00 (0.93 to 1.07) |
| IMD 4 | 8,060 (9.9) | 19.2 | 0.91 (0.85 to 0.97) | 3,120 (9.6) | 24.7 | 0.86 (0.79 to 0.95) |
| IMD 5 (least deprived) | 5,525 (6.8) | 18.8 | 0.90 (0.84 to 0.97) | 1,780 (5.5) | 26.1 | 0.93 (0.83 to 1.04) |
| Comorbid Long Term Condition | | | | | | |
| Type 2 Diabetes | 51,035 (62.5) | 17.5 | 0.66 (0.64 to 0.69) | 15,275 (47.1) | 22.2 | 0.69 (0.66 to 0.73) |
| Type 1 Diabetes | 565 (0.7) | 26.5 | | 415 (1.3) | 25.3 | 0.87 (0.69 to 1.09) |
| Chronic Cardiac Disease | 18,950 (23.2) | 19.9 | 1.12 (1.08 to 1.17) | 4,425 (13.7) | 25.9 | 1.12 (1.04 to 1.20) |
| Learning Disability | 370 (0.5) | 27.0 | 1.11 (0.87 to 1.40) | 200 (0.6) | 32.5 | 1.20 (0.88 to 1.61) |
| Depression | 14,715 (18.0) | 22.8 | 1.15 (1.10 to 1.20) | 5,550 (17.1) | 29.8 | 1.10 (1.03 to 1.17) |
| Serious Mental Illness | 2,135 (2.6) | 26.0 | 1.30 (1.18 to 1.44) | 1,440 (4.4) | 31.6 | 1.18 (1.05 to 1.32) |
| Dementia | 1,905 (2.3) | 20.5 | 1.16 (1.03 to 1.30) | 750 (2.3) | 24.0 | 1.16 (0.97 to 1.38) |
| Asthma | 16,255 (19.9) | 21.1 | 1.06 (1.01 to 1.10) | 5,525 (17.0) | 28.3 | 1.07 (1.00 to 1.14) |
| COPD | 3,150 (3.9) | 20.8 | 1.18 (1.08 to 1.29) | 1,005 (3.1) | 26.4 | 1.20 (1.04 to 1.38) |
| Stroke and TIA | 5,905 (7.2) | 21.1 | 1.31 (1.08 to 1.58) | 2,065 (6.4) | 24.2 | 1.00 (0.90 to 1.11) |

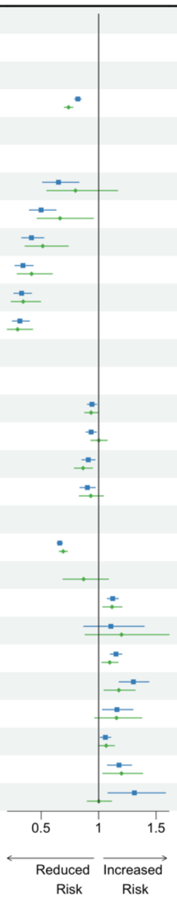

Rapid weight gain: > 0.5 kilograms/metre$^2$/year. N (%): Total number (N) and percentage (%) from each population strata contributing to the analysis. Prevalence (%): percentage who gained weight rapidly. aOR: adjusted Odds Ratio generated in logistic regression models adjusted for age, sex, and Index of Multiple Deprivation. 95% CI: 95% Confidence Interval of aOR. COPD: Chronic Obstructive Pulmonary Disease. TIA: Transient Ischaemic Attack.

Figure S10. Age stratified odds of rapid weight gain amongst adults living in England with hypertension during the COVID-19 pandemic.

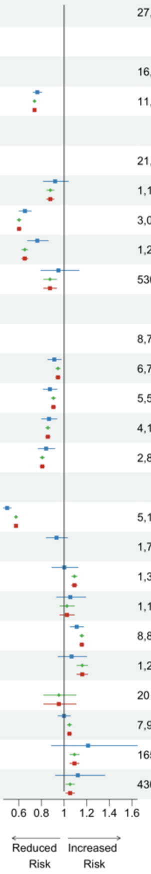

| Population Subgroup | 18-39 years N (%) | 18-39 years Prevalence (%) | 18-39 years aOR (95% CI) | 40-59 years N (%) | 40-59 years Prevalence (%) | 40-59 years aOR (95% CI) | 60-79 years N (%) | 60-79 years Prevalence (%) | 60-79 years aOR (95% CI) |
|---|---|---|---|---|---|---|---|---|---|
| All | 27,960 (100.0) | 39.9 | | 342,510 (100.0) | 30.9 | | 342,510 (100.0) | 30.9 | |
| Sex | | | | | | | | | |
| Female | 16,780 (60.0) | 42.6 | 1 | 159,030 (46.4) | 34.4 | 1 | 159,030 (46.4) | 34.4 | 1 |
| Male | 11,175 (40.0) | 35.7 | 0.76 (0.73 to 0.80) | 183,475 (53.6) | 27.9 | 0.74 (0.73 to 0.75) | 183,475 (53.6) | 27.9 | 0.74 (0.73 to 0.75) |
| Ethnicity | | | | | | | | | |
| White | 21,985 (78.6) | 41.3 | 1 | 279,300 (81.5) | 32.2 | 1 | 279,300 (81.5) | 32.2 | 1 |
| Black | 1,170 (4.2) | 40.2 | 0.92 (0.82 to 1.04) | 15,200 (4.4) | 30.7 | 0.88 (0.85 to 0.91) | 15,200 (4.4) | 30.7 | 0.88 (0.85 to 0.91) |
| South Asian | 3,045 (10.9) | 31.5 | 0.65 (0.60 to 0.71) | 29,830 (8.7) | 22.9 | 0.60 (0.59 to 0.62) | 29,830 (8.7) | 22.9 | 0.60 (0.59 to 0.62) |
| Chinese and Other | 1,225 (4.4) | 34.7 | 0.76 (0.68 to 0.86) | 13,855 (4.0) | 24.0 | 0.65 (0.63 to 0.68) | 13,855 (4.0) | 24.0 | 0.65 (0.63 to 0.68) |
| Mixed | 530 (1.9) | 40.6 | 0.95 (0.80 to 1.13) | 4,325 (1.3) | 30.2 | 0.88 (0.82 to 0.93) | 4,325 (1.3) | 30.2 | 0.88 (0.82 to 0.93) |
| Index of Multiple Deprivation | | | | | | | | | |
| IMD 1 (most deprived) | 8,710 (31.2) | 41.4 | 1 | 89,200 (26.0) | 32.5 | 1 | 89,200 (26.0) | 32.5 | 1 |
| IMD 2 | 6,745 (24.1) | 39.5 | 0.91 (0.86 to 0.97) | 75,800 (22.1) | 31.2 | 0.95 (0.93 to 0.97) | 75,800 (22.1) | 31.2 | 0.95 (0.93 to 0.97) |
| IMD 3 | 5,500 (19.7) | 38.9 | 0.87 (0.82 to 0.94) | 70,515 (20.6) | 30.8 | 0.91 (0.89 to 0.93) | 70,515 (20.6) | 30.8 | 0.91 (0.89 to 0.93) |
| IMD 4 | 4,115 (14.7) | 39.1 | 0.87 (0.80 to 0.94) | 58,755 (17.2) | 30.0 | 0.86 (0.84 to 0.88) | 58,755 (17.2) | 30.0 | 0.86 (0.84 to 0.88) |
| IMD 5 (least deprived) | 2,890 (10.3) | 38.6 | 0.84 (0.77 to 0.92) | 48,240 (14.1) | 28.9 | 0.81 (0.79 to 0.83) | 48,240 (14.1) | 28.9 | 0.81 (0.79 to 0.83) |
| Comorbid Long Term Condition | | | | | | | | | |
| Type 2 Diabetes | 5,100 (18.2) | 26.3 | 0.50 (0.46 to 0.53) | 116,155 (33.9) | 23.0 | 0.58 (0.57 to 0.58) | 116,155 (33.9) | 23.0 | 0.58 (0.57 to 0.58) |
| Type 1 Diabetes | 1,785 (6.4) | 37.8 | 0.93 (0.84 to 1.03) | 7,995 (2.3) | 32.1 | 1.06 (1.01 to NA) | 7,995 (2.3) | 32.1 | 1.06 (1.01 to NA) |
| Chronic Cardiac Disease | 1,305 (4.7) | 39.1 | 1.00 (0.89 to 1.12) | 39,020 (11.4) | 31.8 | 1.09 (1.07 to 1.12) | 39,020 (11.4) | 31.8 | 1.09 (1.07 to 1.12) |
| Learning Disability | 1,150 (4.1) | 40.0 | 1.05 (0.93 to 1.19) | 4,620 (1.3) | 32.4 | 1.03 (0.96 to 1.09) | 4,620 (1.3) | 32.4 | 1.03 (0.96 to 1.09) |
| Depression | 8,875 (31.7) | 42.9 | 1.11 (1.05 to 1.17) | 110,200 (32.2) | 34.4 | 1.16 (1.14 to 1.17) | 110,200 (32.2) | 34.4 | 1.16 (1.14 to 1.17) |
| Serious Mental Illness | 1,230 (4.4) | 40.2 | 1.07 (0.95 to 1.20) | 10,855 (3.2) | 34.2 | 1.16 (1.11 to 1.21) | 10,855 (3.2) | 34.2 | 1.16 (1.11 to 1.21) |
| Dementia | 20 (0.1) | 50.0 | Redacted | 835 (0.2) | 30.5 | 0.95 (0.82 to 1.10) | 835 (0.2) | 30.5 | 0.95 (0.82 to 1.10) |
| Asthma | 7,990 (28.6) | 40.4 | 1.00 (0.95 to 1.05) | 75,145 (21.9) | 32.6 | 1.05 (1.03 to 1.07) | 75,145 (21.9) | 32.6 | 1.05 (1.03 to 1.07) |
| COPD | 165 (0.6) | 45.5 | 1.21 (0.89 to 1.65) | 13,460 (3.9) | 34.2 | 1.09 (1.05 to 1.13) | 13,460 (3.9) | 34.2 | 1.09 (1.05 to 1.13) |
| Stroke and TIA | 430 (1.5) | 43.0 | 1.12 (0.92 to 1.36) | 14,265 (4.2) | 32.1 | 1.05 (1.02 to 1.09) | 14,265 (4.2) | 32.1 | 1.05 (1.02 to 1.09) |

Rapid weight gain: > 0.5 kilograms/metre$^2$/year. N (%): Total number (N) and percentage (%) from each population strata contributing to the analysis. Prevalence (%): percentage who gained weight rapidly. aOR: adjusted Odds Ratio generated in logistic regression models adjusted for sex, ethnicity, and Index of Multiple Deprivation. 95% CI: 95% Confidence Interval of aOR. COPD: Chronic Obstructive Pulmonary Disease. TIA: Transient Ischaemic Attack.
